# Sarilumab treatment of hospitalised patients with severe or critical COVID-19: a multinational, randomised, adaptive, phase 3, double-blind, placebo-controlled trial

**DOI:** 10.1101/2021.02.01.21250769

**Authors:** Francois-Xavier Lescure, Hitoshi Honda, Robert A. Fowler, Jennifer Sloane Lazar, Genming Shi, Peter Wung, Naimish Patel, Owen Hagino

## Abstract

**Background:** Elevated proinflammatory cytokines have been associated with 2019 coronavirus disease (COVID-19) severity. We assessed efficacy and safety of sarilumab, an interleukin-6 receptor inhibitor, in severe (requiring supplemental oxygen by nasal canula or face mask) or critical (requiring greater supplemental oxygen, mechanical ventilation, or extracorporeal support) COVID-19.

**Methods:** This was a 60-day, randomised, double-blind, placebo-controlled, multinational trial in patients hospitalised with laboratory-confirmed severe acute respiratory syndrome coronavirus 2 (SARS-CoV-2) infection and pneumonia, who required oxygen supplementation or intensive care. Patients were randomised 2:2:1 to intravenous sarilumab 400 mg, sarilumab 200 mg, or placebo. The primary endpoint was time to ≥2-point clinical improvement (7-point scale; range: 1 [death] to 7 [not hospitalised]). The key secondary endpoint was proportion of patients alive at day 29. Safety outcomes included adverse events and laboratory assessments. This trial is registered with ClinicalTrials.gov (NCT04327388).

**Findings:** Between March 28 and July 3, 2020, 420 patients were randomised; 416 received treatment (placebo, n=84; sarilumab 200 mg, n=159; sarilumab 400 mg, n=173). At day 29, there were no significant differences in median (95% CI) time to ≥2-point improvement between placebo (12·0 [9·0–15·0] days) and sarilumab groups (200 mg: 10·0 [9·0–12·0] days, p=0.96, log-rank test; 400 mg: 10·0 [9·0–13·0] days, p=0.34) or in proportions of patients alive (placebo, 91·7%; sarilumab 200 mg, 89·9%, p=0·63; sarilumab 400 mg, 91·9%, p=0·85). At day 29, there were numerical, nonsignificant survival differences between sarilumab 400 mg (88%) and placebo (79%; difference +9%, 95% CI −7·7 to 25·5, p=0·25) for critical patients. There were no unexpected safety signals.

**Interpretation:** This trial did not demonstrate efficacy of sarilumab in patients hospitalised with COVID-19 and receiving supplemental oxygen. Adequately powered trials of targeted immunomodulatory therapies assessing survival as a primary endpoint are suggested in patients with critical COVID-19.

**Funding:** Sanofi and Regeneron Pharmaceuticals, Inc.

## Introduction

The emergence of severe acute respiratory syndrome coronavirus 2 (SARS-CoV-2) in December 2019 and the associated coronavirus disease (COVID-19)^1^ has resulted in ≥47 million confirmed infections and ≥1·2 million deaths worldwide as of November 4, 2020. COVID-19-associated pneumonia can rapidly progress to acute respiratory distress syndrome (ARDS), estimated to affect 5–20% of patients with COVID-19.^2-5^ In some patients, COVID-19 may cause damage to additional organs, including heart, brain, kidney, and liver.^6^ In the first several months of the pandemic, there were no treatments with proven efficacy for patients with severe or critical COVID-19; therefore, carefully designed randomised, controlled trials of novel or repurposed medications were, and still are, warranted.

Literature to date supports an association of proinflammatory cytokines with acute, life-threatening respiratory injury observed in patients with COVID-19.^7^ Among these cytokines, interleukin (IL)-6 appears to play a prominent role in the pathogenesis of COVID-19-related ARDS.^8^ Results of a meta-analysis of laboratory findings indicate that 53% of patients with COVID-19 have increased IL-6 levels.^9^ A meta-analysis of 23 clinical trials involving 3400 patients showed that patients with severe COVID-19 had higher levels of IL-6 than those with mild disease, and even higher levels were observed in patients who died.^10^ Two additional meta-analyses^11,12^ and a large, prospective cohort study of patients hospitalised with COVID-19^13^ also demonstrated an association between elevated IL-6 and COVID-19-related mortality. Many of these findings were first published during the early months of the pandemic, suggesting that inhibition of IL-6 signaling may have value as a treatment to manage inflammatory manifestations of COVID-19 pneumonia.

Sarilumab is a human monoclonal antibody that inhibits the binding of IL-6 to its α receptor and is approved for treatment of adults with moderate to severely active rheumatoid arthritis.^14,15^ Because sarilumab inhibits both soluble and membrane-bound forms of IL-6 receptors,^14,16^ potentially suppressing proinflammatory signaling by both pulmonary epithelial and immune cells,^8^ we hypothesised it could reduce the severity of pulmonary complications of COVID-19, including respiratory failure. Here, we report results of a 60-day, randomised, placebo-controlled trial of sarilumab in hospitalised patients with severe to critical COVID-19.

## Methods

### Study design

This study was an adaptive, phase 2/3, multicentre, randomised, double-blinded trial. Because of the uncertainties of assessing treatment efficacy in COVID-19 pneumonia at the time of study design, the initial protocol allowed adaptations such as modification of the provisional phase 3 endpoints, sample size re-estimation before entering phase 3, or closing a dose group while the study remained blinded. Patients were assessed daily while hospitalised until discharge, or death, with a final follow-up on day 60.

The trial was monitored by an external independent data monitoring committee (IDMC) with ongoing access to unblinded clinical data. The protocol was approved by the institutional review boards at each participating hospital and by national ethics committees, as required by local and national regulations. The study was carried out in accordance with the International Conference on Harmonisation Guidelines for Good Clinical Practice and the World Medical Association’s Declaration of Helsinki and its amendments.^17^

### Patients

This study enrolled patients aged 18 years or older at the time of signing informed consent who had been hospitalised for laboratory-confirmed SARS-CoV-2 infection in any specimen within 2 weeks prior to randomisation and with evidence of pneumonia by chest imaging or chest auscultation and no alternative explanation for current clinical presentation. Patients also had to meet criteria for severe disease (defined as administration of supplemental oxygen by nasal cannula, simple face mask, or another similar device) or critical disease (defined as need for supplemental oxygen delivered by nonrebreather mask or high-flow nasal cannula, use of invasive or noninvasive ventilation, or treatment in an intensive care unit). Before participating in the trial, informed consent was obtained from all patients or their legally authorised/appointed representatives, as specified by local law and in compliance with or exceeding ethics committee requirements.

Patients were excluded from the study if they had at least one of the following: in the investigator’s opinion, a low probability of surviving 48 hours or remaining at the investigational site beyond 48 hours, or dysfunction of ≥2 organ systems or need for extracorporeal life support or renal replacement therapy at screening; absolute neutrophil count <2000/mm^3^; aspartate aminotransferase or alanine aminotransferase (ALT) exceeding 5-fold upper limit of normal (ULN) at screening; platelets <50,000/mm^3^ at screening; known active, incompletely treated, suspected or known extrapulmonary tuberculosis; prior or concurrent use of immunosuppressants at screening, including, but not limited to, IL-6 inhibitors or Janus kinase inhibitors within 30 days of baseline; anti-CD20 agents without evidence of B-cell recovery to baseline levels or IL-1 receptor antagonist (anakinra) within 1 week of baseline; abatacept within 8 weeks of baseline; tumor necrosis factor α inhibitors within 2–8 weeks of baseline; alkylating agents, including cyclophosphamide, within 6 months of baseline; cyclosporine, azathioprine, mycophenolate mofetil, leflunomide, or methotrexate within 4 weeks of baseline; or intravenous (IV) immunoglobulin within 5 months of baseline; use of systemic chronic (eg, oral) corticosteroids for a condition not related to COVID-19 at doses higher than prednisone 10 mg/day or equivalent at screening; or suspected or known active systemic bacterial or fungal infections within 4 weeks of screening.

### Randomisation and masking

Eligible patients were randomised (2:2:1) to IV sarilumab 400 mg, sarilumab 200 mg, or placebo according to a central randomisation scheme using permuted blocks of 5 and implemented through an interactive response technology (IRT). Randomisation was stratified by severity of illness (severe or critical) and use of systemic corticosteroids (yes or no).

Patients and investigators remained blinded to patients’ assigned intervention throughout the course of the study. An unblinded pharmacist was responsible for the preparation and dispensation of all study interventions.

### Procedures

Sarilumab 400 mg, sarilumab 200 mg, or placebo were prepared according to instructions provided in the pharmacy manual. After confirming the randomisation number accessed via IRT, the hospital pharmacist added the contents of prefilled syringes (PFS) of sarilumab 200 mg solution for subcutaneous injection supplied by the sponsor into a specified volume of locally sourced 0·9% NaCl solution for IV infusion (two syringes for the 400-mg dose, one syringe for the 200-mg dose, and 0·9% NaCl solution for the placebo dose) to produce an IV bag containing a colourless solution to be administered by blinded hospital staff as a single IV infusion. Patients could have the IV infusion stopped for a safety-related issue, in which case they did not continue with dosing. An option for a second dose existed (within the assigned treatment arm) within 24–48 hours of the first dose, based on the investigator’s benefit-risk assessment (Amended protocol 02; April 8, 2020).

### Clinical and laboratory monitoring

Efficacy assessments included a daily assessment of clinical status until discharge, body temperature (day 1–3: four times a day; day 4–29: twice a day), oxygen administration (day 1–3: four times a day; day 4–29: results recorded as assessed), resting oxygen saturation (SpO2; day 1–3: four times a day; day 4–29: results recorded as assessed), and National Early Warning Score 2 (NEWS2).^18^ Safety procedures and assessments included clinical laboratory testing (performed locally at each hospital), targeted physical examination, and concomitant medication review. Vital signs were recorded daily until discharge. Surveillance testing for bacterial and fungal infection was performed locally, on days 7 and 15. In addition to the positive SARS-CoV-2 result required for inclusion, nasopharyngeal (when feasible) and blood samples were collected at baseline and on days 7, 15, 21, and 29, or on the day of hospital discharge and analysed by the local laboratories and a central laboratory, respectively. Serum IL-6 and other biomarkers were analysed in a central laboratory. Blood samples were also taken for measurement of sarilumab concentration. Other than central laboratory results, all clinical data were entered by investigators at each site into an electronic clinical research form (eCRF) and validated remotely by the sponsor’s monitoring team.

### Outcomes

The primary efficacy endpoint was time from baseline to clinical improvement of ≥2 points on a 7-point ordinal scale, with numerical values defined as follows: 1—Death; 2— Hospitalised, on invasive mechanical ventilation or extracorporeal membrane oxygenation; 3—Hospitalised, on noninvasive ventilation or high-flow oxygen devices; 4—Hospitalised, requiring supplemental oxygen; 5—Hospitalised, not requiring supplemental oxygen – requiring ongoing medical care (COVID-19 related or otherwise); 6—Hospitalised, not requiring supplemental oxygen – no longer requiring ongoing medical care; 7—Not hospitalised. Discharge prior to day 29 was considered as a 2-point improvement. The key secondary efficacy endpoint was the proportion of patients alive at day 29.

The original phase 2 primary endpoint was the time to resolution of fever for at least 48 hours without antipyretics or until discharge (original protocol; March 18, 2020). However, the unanticipated rapid rate of enrolment made the plan to use the phase 2 analysis to select phase 3 efficacy endpoints unfeasible. As a result, the primary and key secondary endpoints for phase 3, as described above, were adopted *a priori* in the Amended protocol 04 (April 8, 2020).

Other secondary efficacy endpoints included differences in time-to-event endpoints by treatment (eg, time to improvement of ≥1 point on the 7-point scale, fever resolution, or discharge from hospital), score changes at specific time points (eg, proportion with 1-point improvement/worsening), and event durations (eg, mechanical ventilation, hospitalisation).

Safety was assessed by investigator reports of adverse events (AEs), serious AEs, AEs of special interest (infusion-related reactions; hypersensitivity reactions; absolute neutrophil count <500/mm^3^ with or without concurrent invasive infection; increase in ALT of at least 3-fold ULN or in excess of 3-fold ULN and at least 2-fold over the baseline level; invasive bacterial or fungal infections of clinical significance with confirmed diagnosis based on the investigator’s assessment with appropriate diagnostic workups and consultations; symptomatic overdose),^19^ and clinical laboratory parameters including lymphocyte count, neutrophil count, and ALT on days 1, 4, 7, 15, 21, and 29 (if still hospitalised).

### Statistical analysis

This study addressed the null hypothesis of no difference in time to ≥2-point improvement on the 7-point scale between a sarilumab dose group and placebo. In sample size determination, approximately 400 patients, randomised 2:2:1, were estimated to provide ≥90% power for pairwise comparison between each sarilumab dose (approximately 160 patients each) and placebo (approximately 80 patients) using a log-rank test of superiority at a two-sided significance level of 0·05. Assumptions included accrual duration of 3 months, each patient being followed up for ≥29 days, and proportions of patients with 2-point improvement at day 15 being 45% for placebo and 70% for sarilumab.

The modified intention-to-treat (mITT) and safety populations included all randomised patients treated with study medication. Primary analysis was planned at day 29 and final analysis at day 60. Analysis of the primary endpoint (mITT) involved a stratified log-rank test with treatment as a fixed factor. Estimation of treatment effect was provided as hazard ratio (HR), generated using a stratified Cox proportional hazards model with treatment as a covariate. Patients without improvement were censored at the last observation time point; patients who took rescue medication in the study without prior improvement were censored at rescue medication start date. Patients who died were deemed no improvement starting from death date. The proportional hazards assumption was assessed by visual inspection of the plot of log(–log(survival)) versus log(survival time) to determine whether curves were parallel among treatments. Analysis of the key secondary endpoint (mITT) involved a Cochran-Mantel-Haenszel test, with estimation of treatment effect reported as the difference in percentage of patients alive at day 29 (sarilumab − placebo). An administrative interim analysis was prespecified for a point in time when approximately 50% of total planned patients (approximately 200) reached day 15. Multiplicity was addressed for the primary and key secondary endpoints for the primary analysis at day 29, by means of hierarchical testing (1. Primary endpoint sarilumab 400 mg *vs* placebo; 2. Key secondary endpoint sarilumab 400 mg *vs* placebo; 3. Primary endpoint sarilumab 200 mg *vs* placebo; 4. Key secondary endpoint sarilumab 200 mg *vs* placebo).

### Amendments made to the original protocol

In total, four amendments were made to the original protocol. Amended protocol 01 (March 26, 2020) implemented clarifications to the original version. Amended protocol 02 (April 8, 2020) implemented changes from phase 2 primary and key secondary endpoints to the phase 3 primary and key secondary endpoints and added an option for a second dose. Amended protocol 03 (April 29, 2020) added an interim analysis when approximately 50% of the total planned number of patients reached day 15, for review by the IDMC and unblinded representatives of sponsor’s senior management, who were not involved in study conduct; clarified the extent to which the sponsor could adapt the study following review of an interim report; and removed the use of vasopressors as an exclusion criterion. Amended protocol 04 (June 11, 2020) closed enrolment into the 200-mg arm following senior management review of interim results and subsequent confirmation by IDMC review, which favoured the 400-mg arm.

## Results

The first patient was screened on March 28, 2020, and the last patient was randomised on July 3, 2020. Last patient – last visit was on September 2, 2020. The study was conducted at 45 sites in Argentina, Brazil, Canada, Chile, France, Germany, Israel, Italy, Japan, Russia, and Spain (table S1). Of 431 patients who were screened, 420 were randomised and 416 received treatment (placebo, n=84; sarilumab 200 mg, n=159; sarilumab 400 mg, n=173) (figure 1).

**Figure 1:**
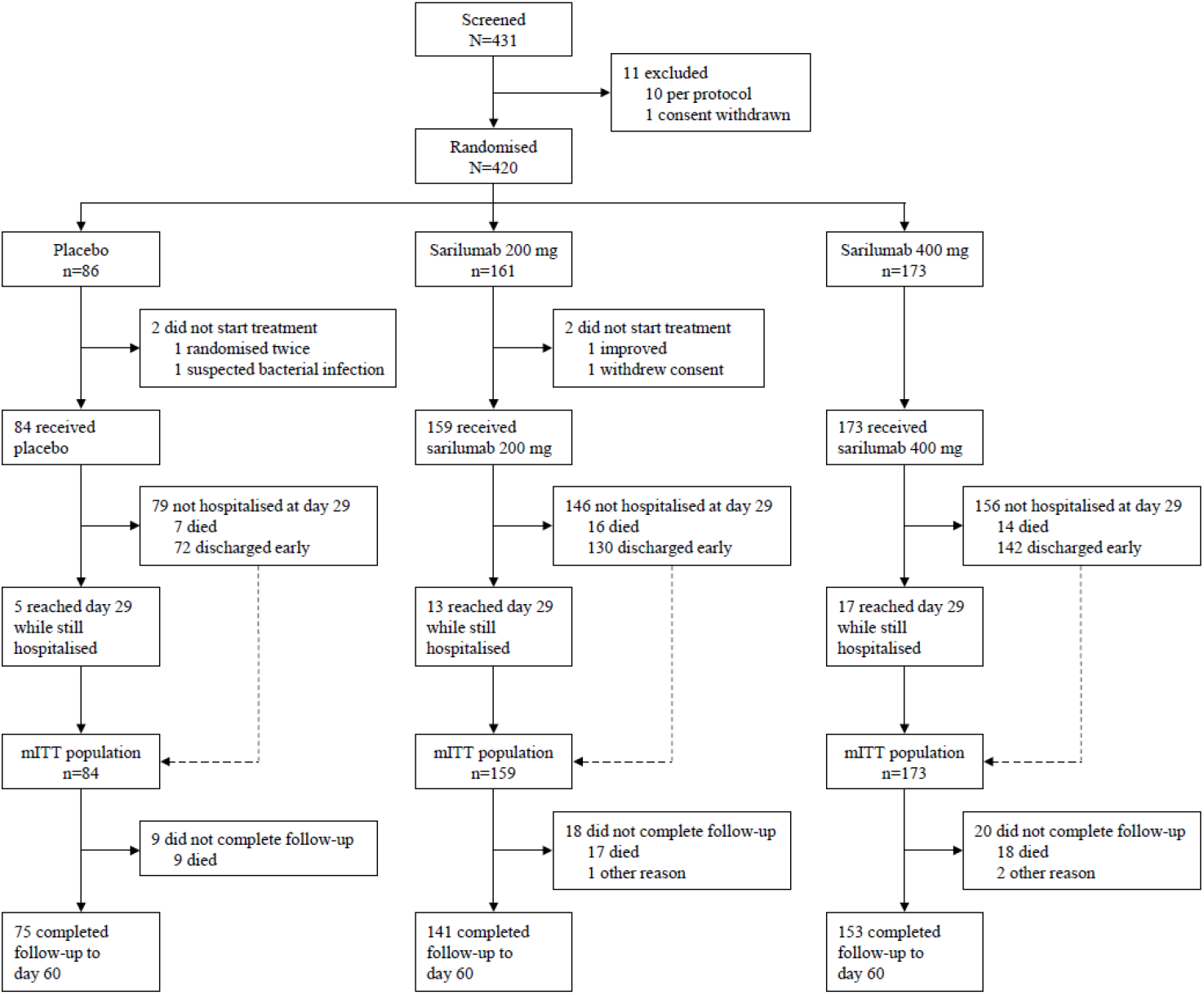
Trial profile. mITT=modified intention-to-treat.

Baseline demographic, clinical, and laboratory characteristics were overall similar among the treatment groups, with exceptions including sex distribution, ferritin concentration, and proportions of patients with fever and obesity (table 1). Median age was 59·0 years (interquartile range [IQR] 50·0–68·0 years) and 37% of participants were women. According to investigator-reported severity, 61% had severe disease and 39% had critical disease. Two (0·5%) patients randomised into the severe disease stratum were recorded in the eCRF as having multisystem organ dysfunction because they required renal replacement therapy.

**Table 1:**
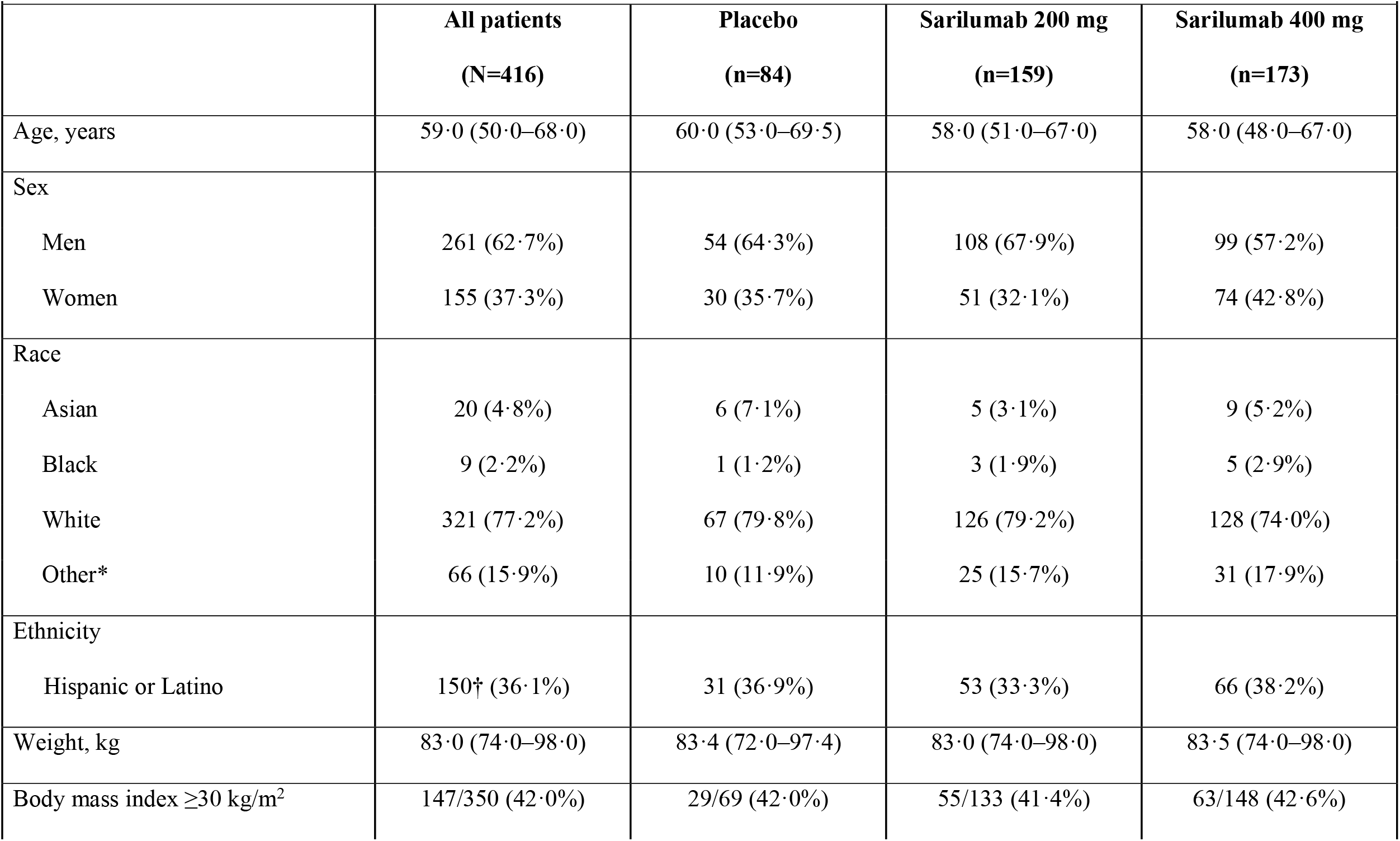
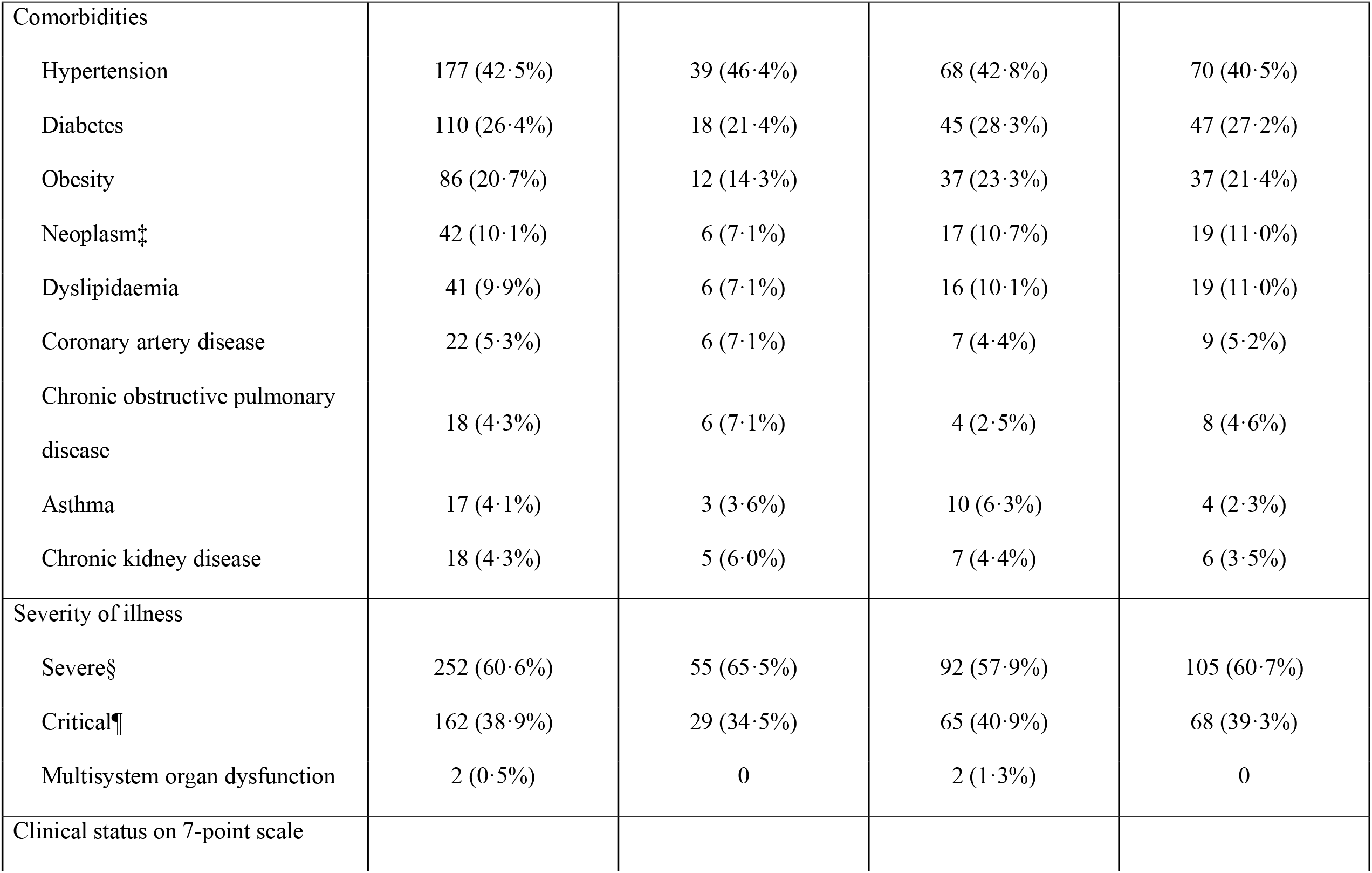
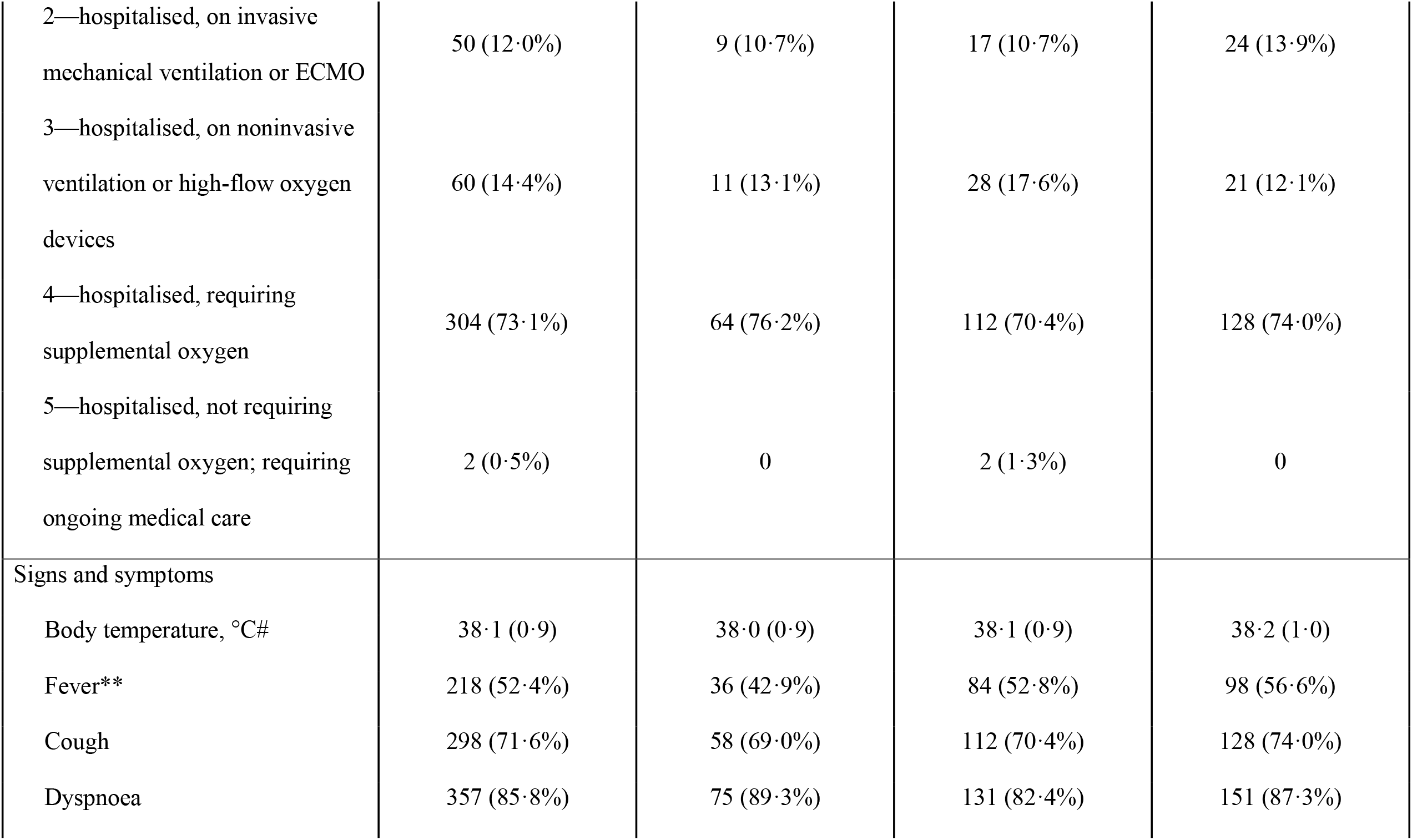
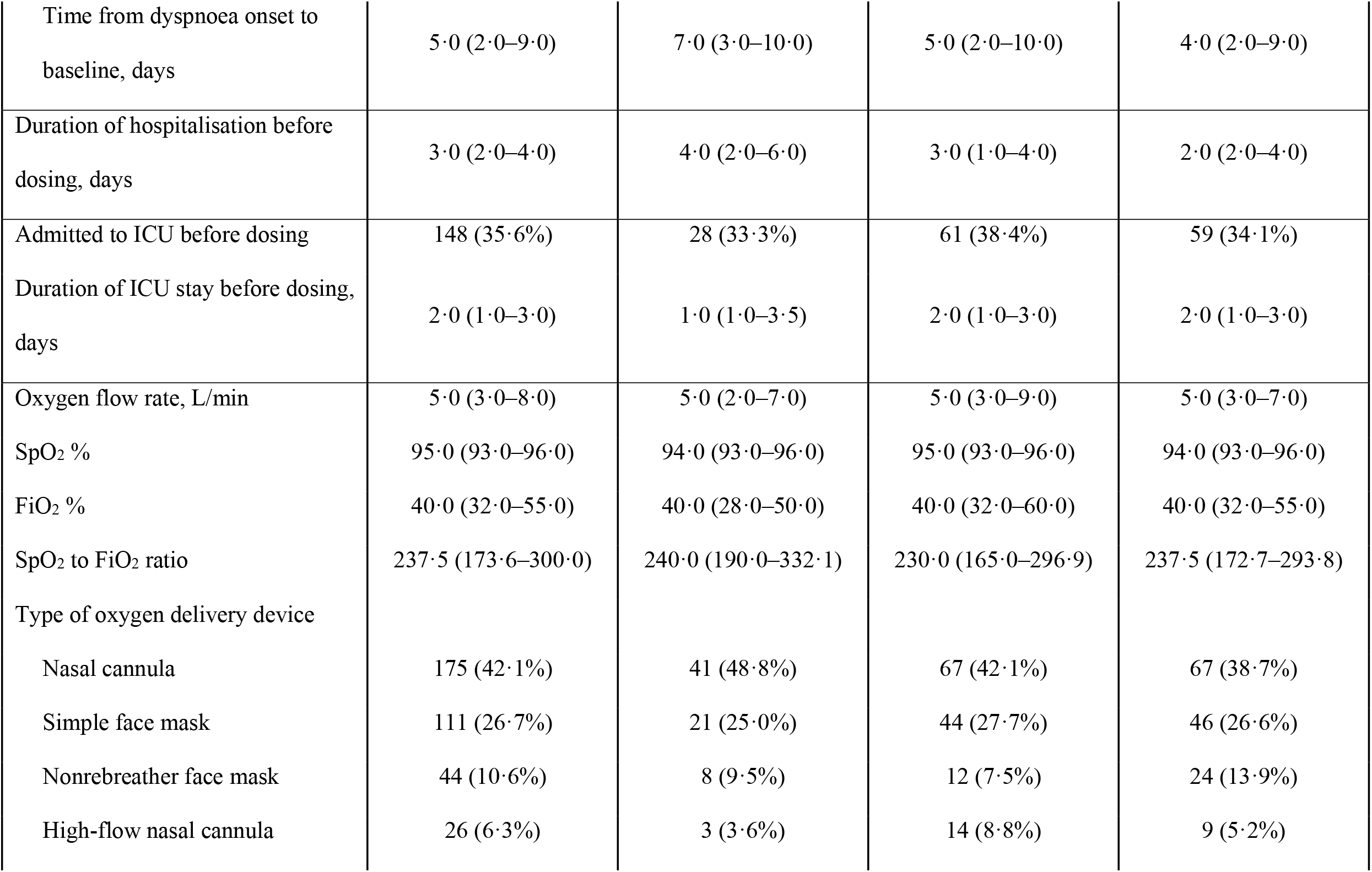
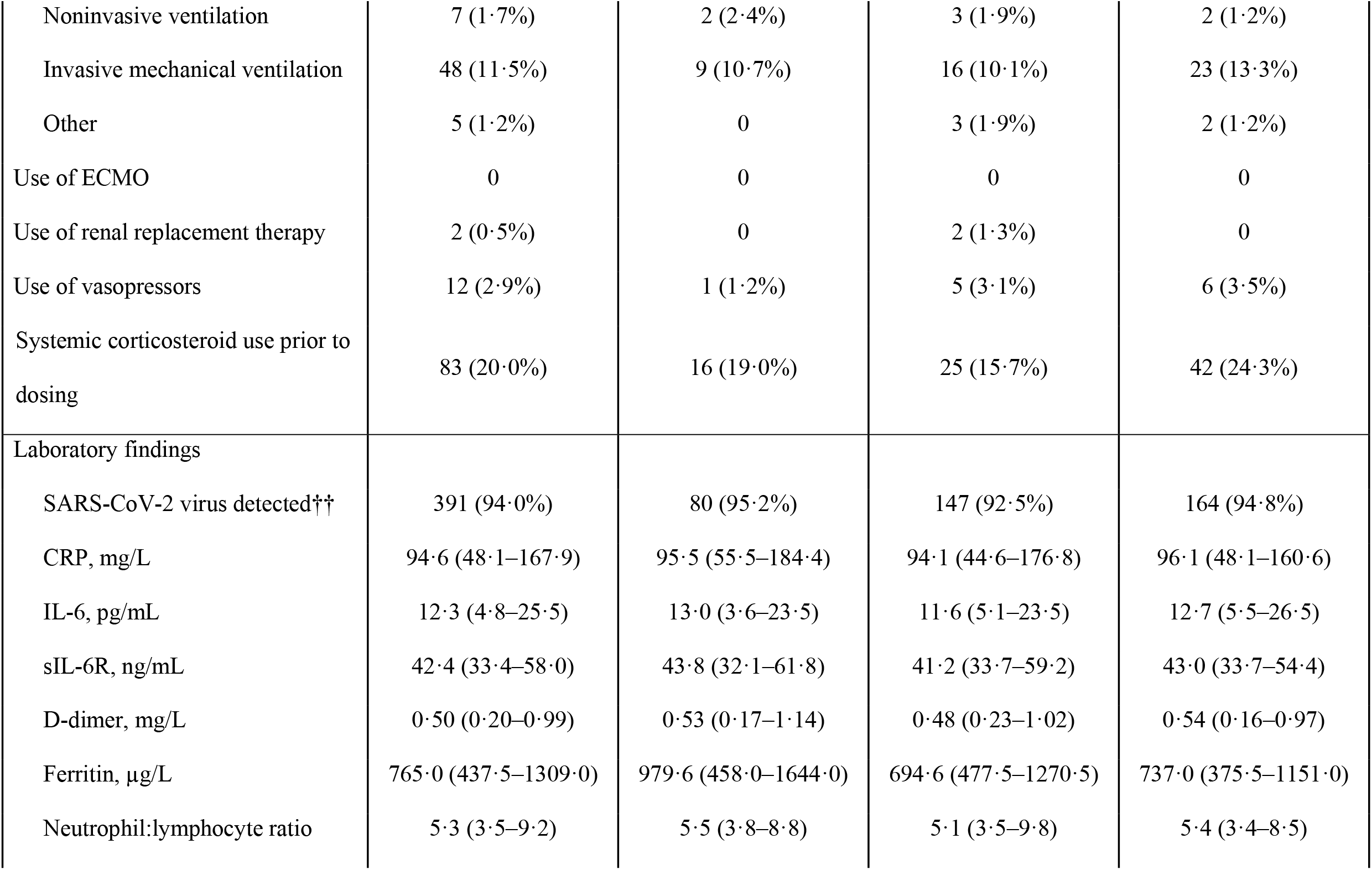
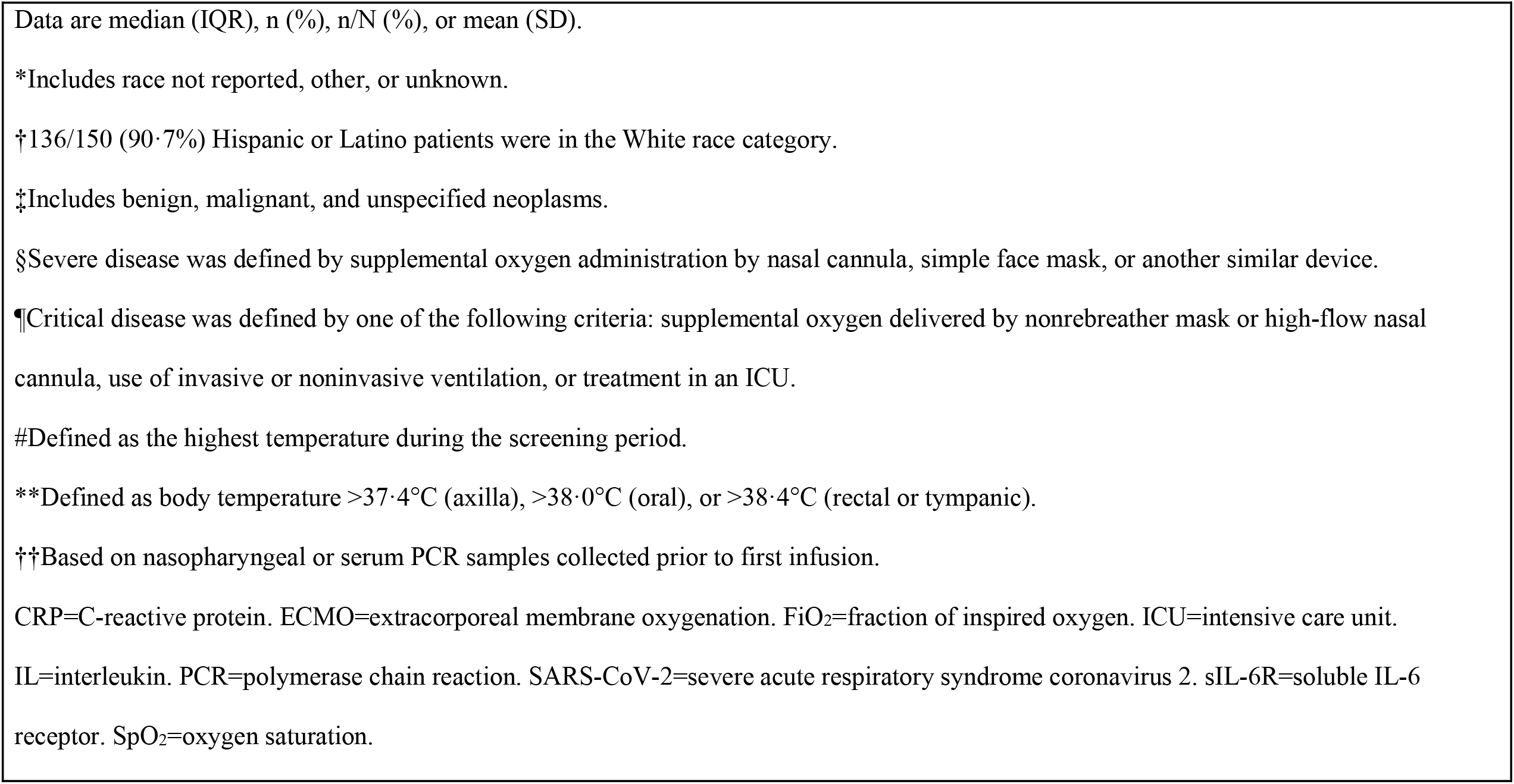
Baseline patient characteristics.

| **Table 1: Baseline patient characteristics.** |  |  |  |  |
| --- | --- | --- | --- | --- |
|  | **All patients**<br>**(N=416)** | **Placebo**<br>**(n=84)** | **Sarilumab 200 mg**<br>**(n=159)** | **Sarilumab 400 mg**<br>**(n=173)** |
| Age, years | 59·0 (50·0–68·0) | 60·0 (53·0–69·5) | 58·0 (51·0–67·0) | 58·0 (48·0–67·0) |
| Sex |  |  |  |  |
| Men | 261 (62·7%) | 54 (64·3%) | 108 (67·9%) | 99 (57·2%) |
| Women | 155 (37·3%) | 30 (35·7%) | 51 (32·1%) | 74 (42·8%) |
| Race |  |  |  |  |
| Asian | 20 (4·8%) | 6 (7·1%) | 5 (3·1%) | 9 (5·2%) |
| Black | 9 (2·2%) | 1 (1·2%) | 3 (1·9%) | 5 (2·9%) |
| White | 321 (77·2%) | 67 (79·8%) | 126 (79·2%) | 128 (74·0%) |
| Other* | 66 (15·9%) | 10 (11·9%) | 25 (15·7%) | 31 (17·9%) |
| Ethnicity |  |  |  |  |
| Hispanic or Latino | 150† (36·1%) | 31 (36·9%) | 53 (33·3%) | 66 (38·2%) |
| Weight, kg | 83·0 (74·0–98·0) | 83·4 (72·0–97·4) | 83·0 (74·0–98·0) | 83·5 (74·0–98·0) |
| Body mass index $\geq 30$ kg/m<sup>2</sup> | 147/350 (42·0%) | 29/69 (42·0%) | 55/133 (41·4%) | 63/148 (42·6%) |
| Comorbidities |  |  |  |  |
| Hypertension | 177 (42·5%) | 39 (46·4%) | 68 (42·8%) | 70 (40·5%) |
| Diabetes | 110 (26·4%) | 18 (21·4%) | 45 (28·3%) | 47 (27·2%) |
| Obesity | 86 (20·7%) | 12 (14·3%) | 37 (23·3%) | 37 (21·4%) |
| Neoplasm‡ | 42 (10·1%) | 6 (7·1%) | 17 (10·7%) | 19 (11·0%) |
| Dyslipidaemia | 41 (9·9%) | 6 (7·1%) | 16 (10·1%) | 19 (11·0%) |
| Coronary artery disease | 22 (5·3%) | 6 (7·1%) | 7 (4·4%) | 9 (5·2%) |
| Chronic obstructive pulmonary disease | 18 (4·3%) | 6 (7·1%) | 4 (2·5%) | 8 (4·6%) |
| Asthma | 17 (4·1%) | 3 (3·6%) | 10 (6·3%) | 4 (2·3%) |
| Chronic kidney disease | 18 (4·3%) | 5 (6·0%) | 7 (4·4%) | 6 (3·5%) |
| Severity of illness |  |  |  |  |
| Severe§ | 252 (60·6%) | 55 (65·5%) | 92 (57·9%) | 105 (60·7%) |
| Critical¶ | 162 (38·9%) | 29 (34·5%) | 65 (40·9%) | 68 (39·3%) |
| Multisystem organ dysfunction | 2 (0·5%) | 0 | 2 (1·3%) | 0 |
| Clinical status on 7-point scale |  |  |  |  |
| 2—hospitalised, on invasive mechanical ventilation or ECMO | 50 (12·0%) | 9 (10·7%) | 17 (10·7%) | 24 (13·9%) |
| 3—hospitalised, on noninvasive ventilation or high-flow oxygen devices | 60 (14·4%) | 11 (13·1%) | 28 (17·6%) | 21 (12·1%) |
| 4—hospitalised, requiring supplemental oxygen | 304 (73·1%) | 64 (76·2%) | 112 (70·4%) | 128 (74·0%) |
| 5—hospitalised, not requiring supplemental oxygen; requiring ongoing medical care | 2 (0·5%) | 0 | 2 (1·3%) | 0 |
| Signs and symptoms |  |  |  |  |
| Body temperature, °C# | 38·1 (0·9) | 38·0 (0·9) | 38·1 (0·9) | 38·2 (1·0) |
| Fever** | 218 (52·4%) | 36 (42·9%) | 84 (52·8%) | 98 (56·6%) |
| Cough | 298 (71·6%) | 58 (69·0%) | 112 (70·4%) | 128 (74·0%) |
| Dyspnoea | 357 (85·8%) | 75 (89·3%) | 131 (82·4%) | 151 (87·3%) |
| Time from dyspnoea onset to baseline, days | 5·0 (2·0–9·0) | 7·0 (3·0–10·0) | 5·0 (2·0–10·0) | 4·0 (2·0–9·0) |
| Duration of hospitalisation before dosing, days | 3·0 (2·0–4·0) | 4·0 (2·0–6·0) | 3·0 (1·0–4·0) | 2·0 (2·0–4·0) |
| Admitted to ICU before dosing | 148 (35·6%) | 28 (33·3%) | 61 (38·4%) | 59 (34·1%) |
| Duration of ICU stay before dosing, days | 2·0 (1·0–3·0) | 1·0 (1·0–3·5) | 2·0 (1·0–3·0) | 2·0 (1·0–3·0) |
| Oxygen flow rate, L/min | 5·0 (3·0–8·0) | 5·0 (2·0–7·0) | 5·0 (3·0–9·0) | 5·0 (3·0–7·0) |
| SpO<sub>2</sub> % | 95·0 (93·0–96·0) | 94·0 (93·0–96·0) | 95·0 (93·0–96·0) | 94·0 (93·0–96·0) |
| FiO<sub>2</sub> % | 40·0 (32·0–55·0) | 40·0 (28·0–50·0) | 40·0 (32·0–60·0) | 40·0 (32·0–55·0) |
| SpO<sub>2</sub> to FiO<sub>2</sub> ratio | 237·5 (173·6–300·0) | 240·0 (190·0–332·1) | 230·0 (165·0–296·9) | 237·5 (172·7–293·8) |
| Type of oxygen delivery device |  |  |  |  |
| Nasal cannula | 175 (42·1%) | 41 (48·8%) | 67 (42·1%) | 67 (38·7%) |
| Simple face mask | 111 (26·7%) | 21 (25·0%) | 44 (27·7%) | 46 (26·6%) |
| Nonrebreather face mask | 44 (10·6%) | 8 (9·5%) | 12 (7·5%) | 24 (13·9%) |
| High-flow nasal cannula | 26 (6·3%) | 3 (3·6%) | 14 (8·8%) | 9 (5·2%) |
| Noninvasive ventilation | 7 (1·7%) | 2 (2·4%) | 3 (1·9%) | 2 (1·2%) |
| Invasive mechanical ventilation | 48 (11·5%) | 9 (10·7%) | 16 (10·1%) | 23 (13·3%) |
| Other | 5 (1·2%) | 0 | 3 (1·9%) | 2 (1·2%) |
| Use of ECMO | 0 | 0 | 0 | 0 |
| Use of renal replacement therapy | 2 (0·5%) | 0 | 2 (1·3%) | 0 |
| Use of vasopressors | 12 (2·9%) | 1 (1·2%) | 5 (3·1%) | 6 (3·5%) |
| Systemic corticosteroid use prior to dosing | 83 (20·0%) | 16 (19·0%) | 25 (15·7%) | 42 (24·3%) |
| Laboratory findings |  |  |  |  |
| SARS-CoV-2 virus detected†† | 391 (94·0%) | 80 (95·2%) | 147 (92·5%) | 164 (94·8%) |
| CRP, mg/L | 94·6 (48·1–167·9) | 95·5 (55·5–184·4) | 94·1 (44·6–176·8) | 96·1 (48·1–160·6) |
| IL-6, pg/mL | 12·3 (4·8–25·5) | 13·0 (3·6–23·5) | 11·6 (5·1–23·5) | 12·7 (5·5–26·5) |
| sIL-6R, ng/mL | 42·4 (33·4–58·0) | 43·8 (32·1–61·8) | 41·2 (33·7–59·2) | 43·0 (33·7–54·4) |
| D-dimer, mg/L | 0·50 (0·20–0·99) | 0·53 (0·17–1·14) | 0·48 (0·23–1·02) | 0·54 (0·16–0·97) |
| Ferritin, µg/L | 765·0 (437·5–1309·0) | 979·6 (458·0–1644·0) | 694·6 (477·5–1270·5) | 737·0 (375·5–1151·0) |
| Neutrophil:lymphocyte ratio | 5·3 (3·5–9·2) | 5·5 (3·8–8·8) | 5·1 (3·5–9·8) | 5·4 (3·4–8·5) |

Data are median (IQR), n (%), n/N (%), or mean (SD).
\*Includes race not reported, other, or unknown.
†136/150 (90·7%) Hispanic or Latino patients were in the White race category.
‡Includes benign, malignant, and unspecified neoplasms.
§Severe disease was defined by supplemental oxygen administration by nasal cannula, simple face mask, or another similar device.
¶Critical disease was defined by one of the following criteria: supplemental oxygen delivered by nonrebreather mask or high-flow nasal cannula, use of invasive or noninvasive ventilation, or treatment in an ICU.

Fever was reported in 52% of patients. Median duration of hospitalisation before dosing was 3·0 days (IQR 2·0–4·0 days). Use of systemic corticosteroids (including dexamethasone), antiviral medications, antibacterial medications (including azithromycin), and hydroxychloroquine/chloroquine prior to, prior to and during, and after first infusion of study medication did not differ substantially across treatment arms (table S2).

For the primary endpoint of time to improvement of ≥2 points on a 7-point clinical assessment scale, no significant difference was observed between sarilumab doses and placebo up to day 29 (figure 2). Although the between-group differences were not significant, median time to improvement was 2 days longer in the placebo group compared with the sarilumab groups (12 days *vs* 10 days) (table 2). In addition, no significant differences were observed in the proportions of patients alive at day 29 (placebo, 91·7%; sarilumab 200 mg, 89·9%; sarilumab 400 mg, 91·9%) (table 2).

**Table 2:**
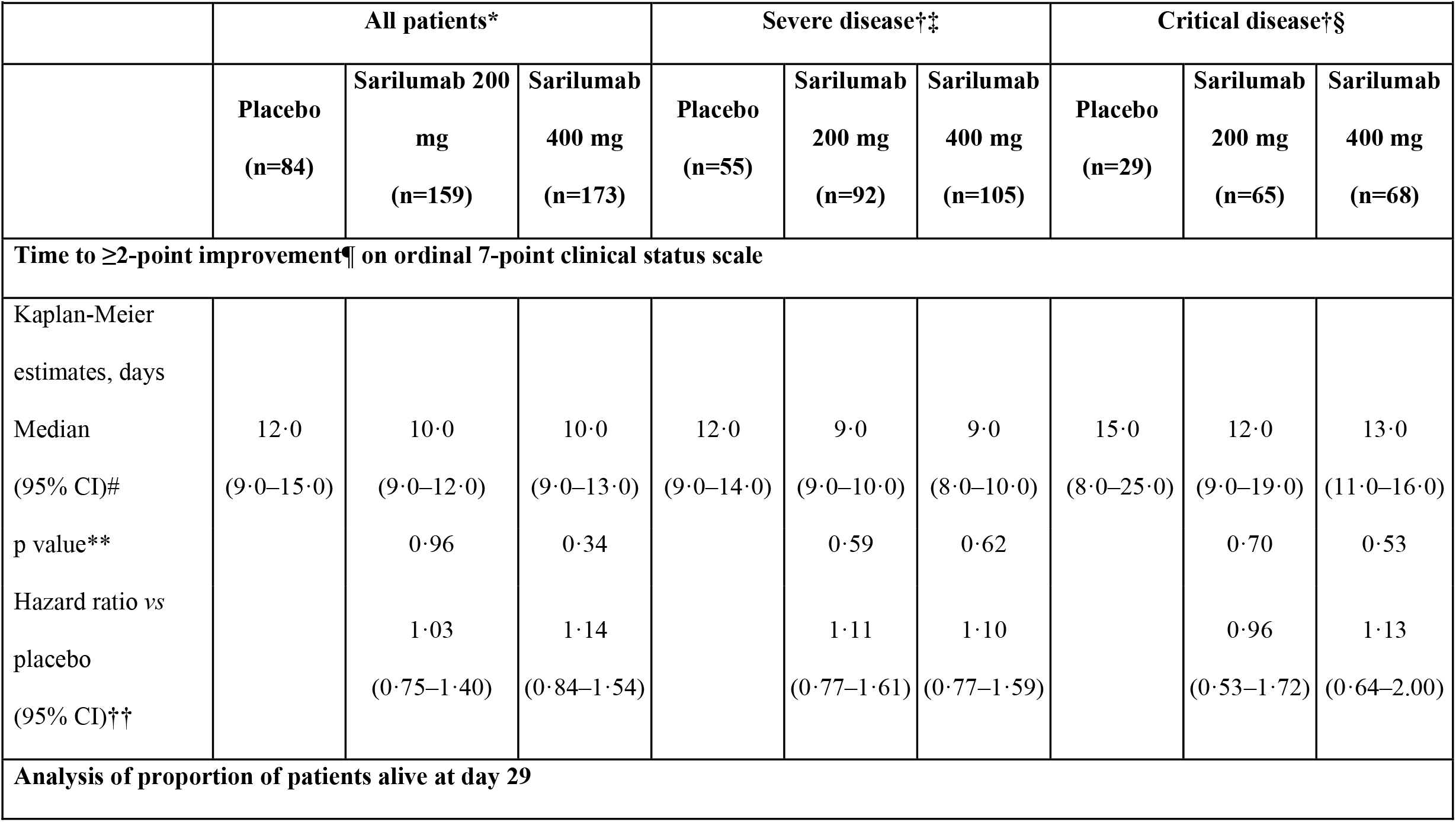
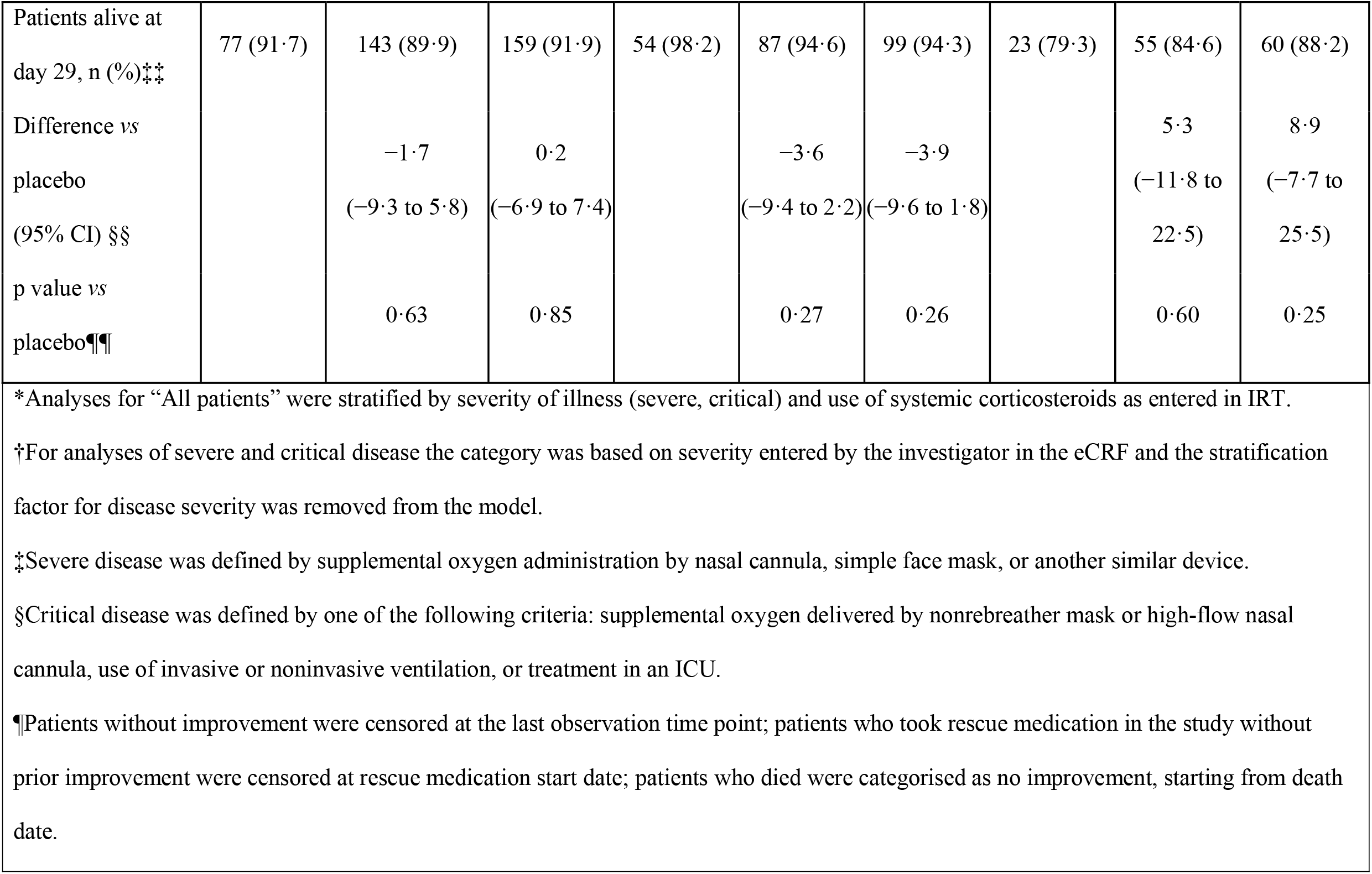
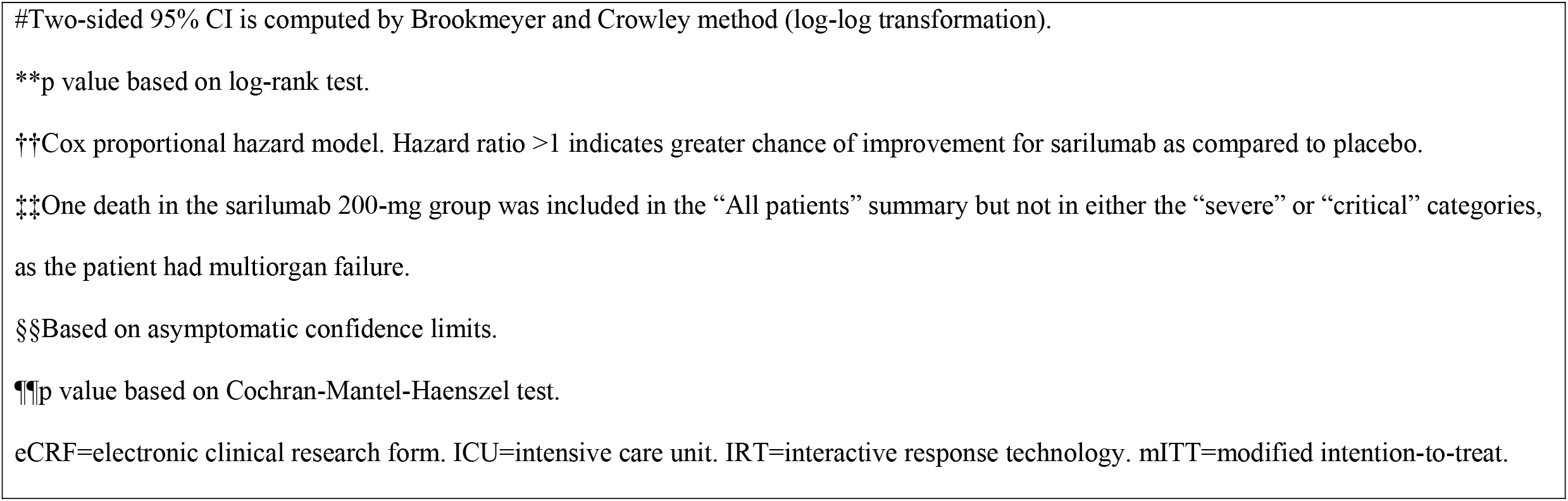
Summary of primary and key secondary endpoints according to investigator-reported disease severity (mITT population) (Day 29 analysis)

**Figure 2:**
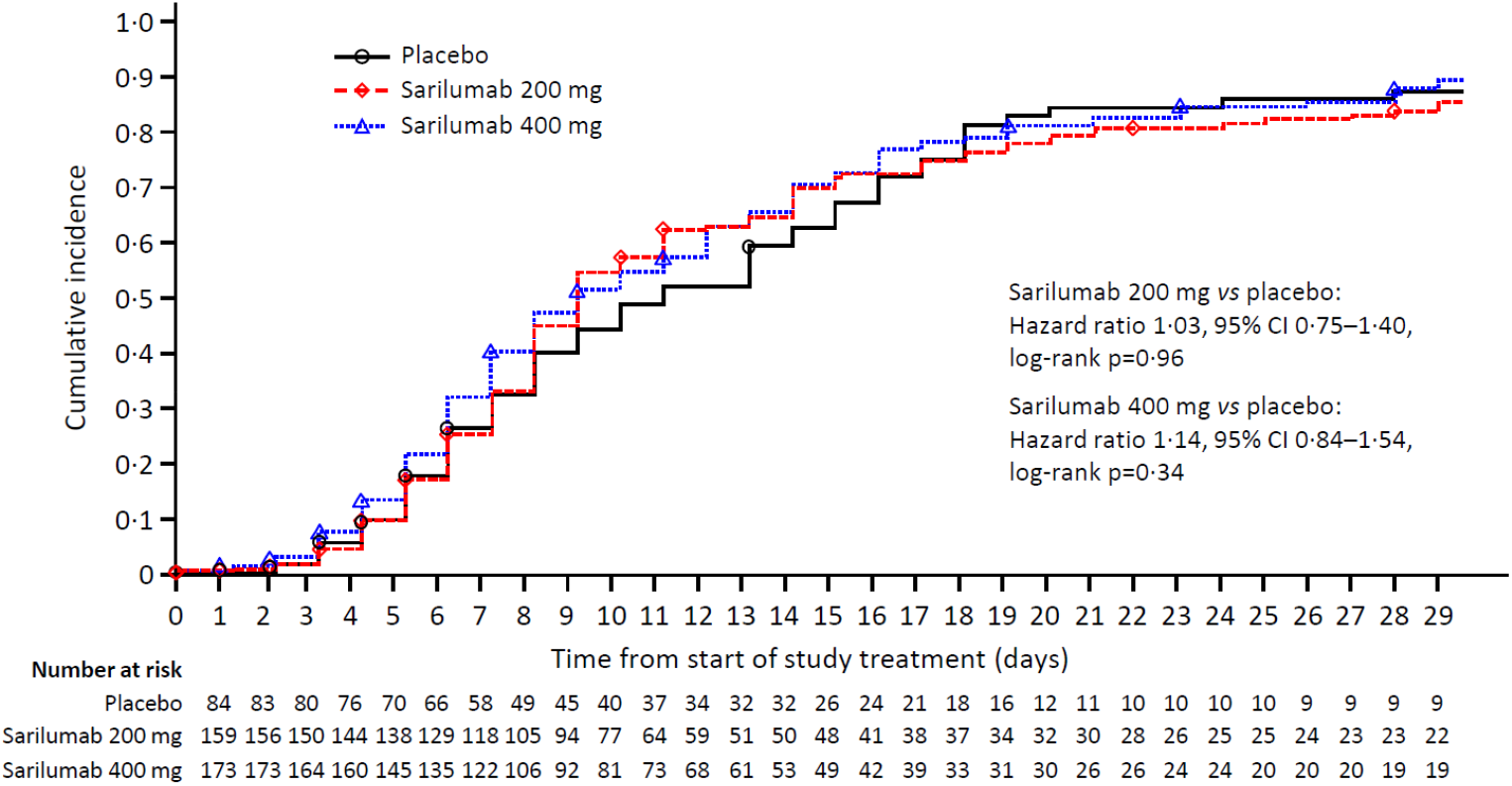
Primary endpoint: time to improvement of ≥2 points in clinical status of assessment from baseline on a 7-point ordinal scale (Kaplan–Maier curves; day 29 analysis). A higher Kaplan–Meier estimate of cumulative incidence indicates a higher proportion of patients with an improvement of at least 2 points in clinical status.

The proportions of patients discharged due to recovery by day 29 were 83·3% (placebo), 79·2% (sarilumab 200 mg), and 79·2% (sarilumab 400 mg) and the percentages of patients alive at day 60 were 89·3%, 89·3%, and 89·6%, respectively (table S3). Additional secondary endpoints related to fever, oxygenation, and hospital status are presented in the supplementary materials (table S3).

Prespecified analysis of day 29 data showed a numerical but nonsignificant difference in survival between sarilumab 400 mg (88%) and placebo (79%; difference +9%, 95% CI −7·7 to 25·5, p=0·25) for the sickest patients in this trial, those with critical disease (table 2).

In patients with either severe or critical disease, differences between sarilumab 400 mg and placebo on the 7-point clinical status scale were larger during the first 2 weeks of treatment than during the second 2 weeks (figure 2). The time-concentration plot of sarilumab concentration following IV infusion showed an initially rapid decline over the first 4 days and a slower decline from day 7 onward, with nearly complete elimination by day 21 even among patients who received two doses of 400 mg within the first 48 hours (figure S2A).

Changes in C-reactive protein (CRP) and neutrophil counts were considered pharmacodynamic markers of systemic IL-6 signaling inhibition. The decline in mean CRP was steeper in the sarilumab arms than in the placebo arm, with a rebound at day 7 in the 200-mg arm and day 15 in the 400-mg arm (figure S2B). As expected with IL-6 receptor inhibition, neutrophil counts were decreased in the sarilumab arms and lower for a longer period of time with the 400-mg dose than the 200-mg dose, but again appeared to increase after day 4 in the 200-mg arm and after day 15 in the 400-mg arm. In the placebo arm, neutrophil counts were stable through day 7 but were higher at day 15 (figure S2E). Neutrophil/lymphocyte ratios (figure S2F) and D-dimer (figure S2G) concentrations did not appear to be influenced by sarilumab concentration, and mean ALT elevation only appeared higher than placebo in the sarilumab arms at day 7 (figure S2H). The concentration-time plots for IL-6 (figure S2C) and soluble IL-6 receptor (sIL-6R) (figure S2D) were consistent with what has been previously reported for sarilumab following subcutaneous injection.^20^ Mean sIL-6R concentration in the placebo arm remained low up to day 29.

The rates of treatment-emergent AEs, infections (including serious infections), and treatment-emergent AEs leading to death were similar among the treatment groups (table 3). In sarilumab-treated patients, the types of AEs of special interest were generally consistent with the established safety profile of sarilumab and the IV route of administration and occurred more frequently than in the placebo group.

**Table 3:** Summary of AEs.

| **Table 3: Summary of AEs.** |  |  |  |
| --- | --- | --- | --- |
| **Patients with $\geq 1$ event** | **Placebo**<br>**(n=84)** | **Sarilumab 200 mg**<br>**(n=159)** | **Sarilumab 400 mg**<br>**(n=173)** |
| Any treatment-emergent AE | 55 (65.5) | 103 (64.8) | 121 (69.9) |
| Any serious treatment-emergent AE | 20 (23.8) | 42 (26.4) | 51 (29.5) |
| Any serious infection | 10 (11.9) | 18 (11.3) | 22 (12.7) |
| Pneumonia | 0 | 1 (0.6) | 6 (3.5) |
| COVID-19 pneumonia | 2 (2.4) | 11 (6.9) | 4 (2.3) |
| Pneumonia bacterial | 1 (1.2) | 1 (0.6) | 3 (1.7) |
| Any treatment-emergent AE leading to death | 9 (10.7) | 17 (10.7) | 18 (10.4) |
| Any AE of special interest | 18 (21.4) | 53 (33.3) | 76 (43.9) |
| ALT increase | 16 (19.0) | 48 (30.2) | 55 (31.8) |
| Invasive bacterial or fungal infection | 3 (3.6) | 8 (5.0) | 15 (8.7) |
| Grade $\geq 2$ hypersensitivity reaction | 0 | 1 (0.6) | 7 (4.0) |
| Grade 4 neutropenia | 0 | 3 (1.9) | 6 (3.5) |
| Grade $\geq 2$ infusion-related reaction | 0 | 1 (0·6) | 6 (3·5) |
| Data are n (%).<br>AE=adverse event. ALT=alanine aminotransferase. COVID-19=coronavirus disease 2019. |  |  |  |

Because standards of care for hospitalised patients with COVID-19 evolved over the course of the trial, in a *post hoc* analysis the proportions of patients initiating or continuing selected medications were plotted by week of study conduct (figure S3). Use of systemic corticosteroids appeared to wane during the first 6 weeks of study conduct, then increased to a peak usage in 70% of patients 13 weeks after the first randomised patient started receiving a corticosteroid (figure S3A). This uptick in corticosteroid usage coincided with increased enrolment of patients with critical disease. Over the course of the study, initiation of systemic corticosteroids did not appear to be related to treatment arm (figure S3A). Use of antiviral medications (figure S3C), hydroxychloroquine/chloroquine (figure S3D), and combinations (figure S4) declined over the course of the trial. For the medications of interest, changes in background therapy appeared balanced across the treatment arms. In subgroup analyses (not presented), no significant interactions between use of systemic corticosteroids, antiviral medications, antibiotic medications, or hydroxychloroquine/chloroquine and time to clinical improvement ≥2 points or survival at day 29 were identified. Only two patients each were treated with remdesivir or convalescent plasma during the trial.

## Discussion

In this multinational, randomised, placebo-controlled study of patients with severe or critical COVID-19 who were receiving the local standard of care, there was no demonstrated benefit of IV sarilumab over placebo. The treatment groups had similar rates of serious infections and AEs leading to death, and types of AEs were consistent with prior clinical trial experience with sarilumab.^15^ No new safety signals for sarilumab were observed in these patients with COVID-19.

There are several potential reasons sarilumab was not effective as a treatment for COVID-19 in this clinical trial. First, IL-6 suppression alone may be insufficient to quell the inflammatory phase of the disease.^21^ Open-label studies in patients with COVID-19 suggested clinical improvement with tocilizumab, another IL-6 inhibitor.^20,22,23^ However, in recently published randomized trials, which also included patients hospitalised with COVID-19 pneumonia, but on average less severely affected than ours,^24-26^ tocilizumab failed to reduce disease severity at day 4 or mortality at day 28,^24^ a clinical worsening at day 14,^25^ or time to intubation or death.^26^

Second, we did not select patients based on commonly available biological and clinical markers of inflammation (eg, elevated CRP) or worsening prognosis (eg, neutrophil counts or uncontrollable fever); consequently, we may not have included a sufficient number of patients for whom immunomodulatory therapy would have been appropriate. Additionally, we may not have chosen an optimal time in the disease course of COVID-19 to administer sarilumab.

Third, immunomodulation may only be beneficial for the most serious cases of COVID-19. Results of a large, open-label, controlled trial of dexamethasone (n=2104) versus usual care (n=4321) for hospitalised patients with COVID-19 suggest the magnitude of survival benefit is related to intensity of respiratory support.^27^ In our study, a numerical difference in survival favouring sarilumab was only seen in the patients who required intensive respiratory support (oxygen by nonrebreather mask or high-flow nasal cannula, use of invasive or noninvasive ventilation), or treatment in an intensive care unit. The differences in treatment response between patients with severe disease and critical disease may be qualitatively reflected in the different evolution of clinical status over the course of the trial; ie, earlier improvement in the sarilumab arms among severely ill patients up to day 15, and greater proportions of patients surviving after day 15 among critically ill patients (figure 3). Kaplan-Meier time-to-event curves up to day 60 also suggest more rapid improvement and earlier discharge due to improvement in the sarilumab arms than the placebo arm among the patients with severe disease; and higher probability of survival among sarilumab-treated patients than placebo-treated in the critically ill group (table 2, figure S1).

**Figure 3:**
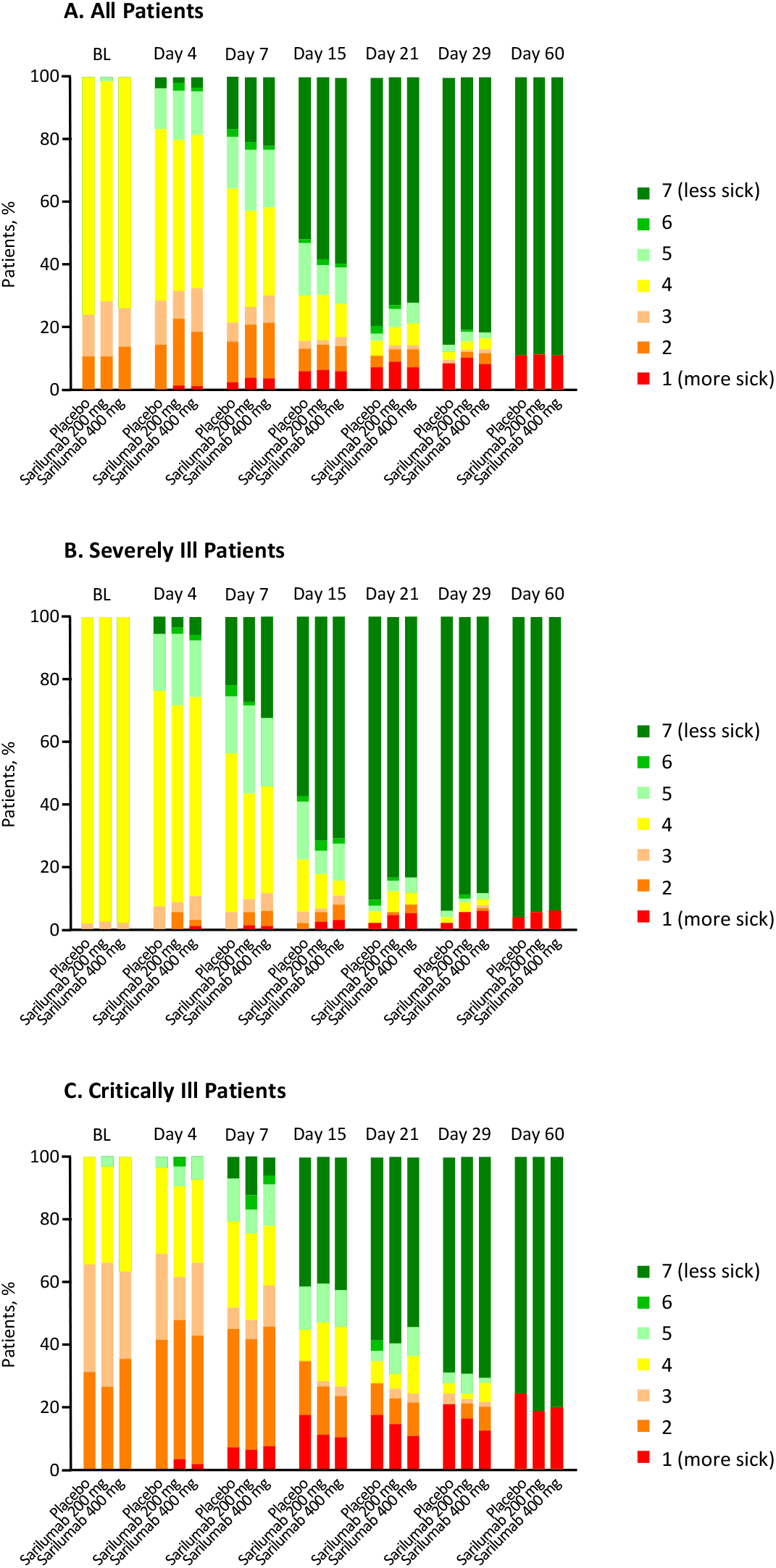
Proportions in each 7-point ordinal scale category over time among all patients (A), severely ill patients (B), and critically ill patients (C). Scores: 1—Death; 2—Hospitalised, on invasive mechanical ventilation or extracorporeal membrane oxygenation; 3—Hospitalised, on noninvasive ventilation or high-flow oxygen devices; 4—Hospitalised, requiring supplemental oxygen; 5—Hospitalised, not requiring supplemental oxygen – requiring ongoing medical care (COVID-19 related or otherwise); 6—Hospitalised, not requiring supplemental oxygen – no longer requiring ongoing medical care; 7—Not hospitalised. ECMO=extracorporeal membrane oxygenation.

Fourth, frequent use of systemic corticosteroids may have reduced the differences between the investigational treatment and the placebo control arms. Over 60% of patients in the trial received at least one dose of systemic corticosteroids before, during, and/or after infusion of the study medication (table S2) and the frequency of systemic corticosteroid varied during the conduct of the study (figure S3).

Fifth, this study may have been underpowered. Significant efficacy results for remdesivir compared with placebo required 1062 patients to show a difference in median recovery time of 5 days^28^ and >1000 patients to show a 1-day difference in median time to recovery of baricitinib on top of remdesivir.^29^

Sixth, a single IV administration of sarilumab 400 mg may be insufficient to control the inflammatory phase of COVID-19 beyond 14 days, as suggested by the reduction in sarilumab concentration between day 7 and day 14 and subsequent rebound in CRP concentration and neutrophil counts after day 15. Alternatively, the IV route of administration, although theoretically advantageous, may not have resulted in a time-concentration profile suited for COVID-19.

Lastly, the efficacy endpoints chosen may have been insufficiently sensitive for the wide range of patients included in the trial. Additionally, an ordinal clinical status scale based on intensity of respiratory support may be too crude to measure treatment effects in patients with an acute systemic disease involving multiple organ systems.

Despite these limitations, survival at day 29 was possibly higher by 9% in the sarilumab arms than in the placebo arm for patients who required noninvasive or invasive mechanical ventilation or ECMO at baseline. Therefore, we think the results of this study do not exclude the possibility of a benefit from targeted immunomodulation in hospitalised patients with COVID-19 pneumonia with critical illness and suggest that subsequent randomised trials of targeted immunomodulatory therapies in this disease focus on critically ill patients and are adequately powered to assess survival as a primary endpoint.

## Supporting information

CONSORT checklist

## Data Availability

Qualified researchers may request access to patient-level data and related study documents including the clinical study report, study protocol with any amendments, blank case report form, statistical analysis plan, and data set specifications. Patient-level data will be anonymised and study documents will be redacted to protect the privacy of trial participants. Further details on Sanofi's data-sharing criteria, eligible studies, and process for requesting access can be found at: https://www.clinicalstudydatarequest.com

## Contributors

F-XL, JSL, GS, and OH had full access to all of the data in the study and take responsibility for the integrity of the data and the accuracy of the data analysis. JSL, GS, PW, and OH provided input on the trial design. F-XL, HH, and RF were responsible for acquisition and interpretation of data. F-XL and OH drafted the manuscript. F-XL, HH, RF, NP, and OH critically revised the manuscript. GS contributed to statistical analysis. F-XL, HH, RF, JSL, and PW contributed to conducting the trial. All authors reviewed and approved the final version of the manuscript.

## Acknowledgments

The authors and Sanofi thank the patients for their participation in the trial, as well as the investigators. Special thanks to Meng Zhang, Christine Xu, and Evelyne Dombrecht of Sanofi for their expertise, dedication, and leadership of a remarkable team, and to the members of the IDMC (Kevin Winthrop MD, MPH; Victor Ortega MD, PhD, ATSF; Mitchell Levy MD, Steve Dahlberg MS) and the independent Statistical Data Analysis Center (Department of Biostatistics and Medical Informatics, University of Wisconsin–Madison) for their diligence and equipoise under extraordinary circumstances. This study was funded by Sanofi (Paris, France) and Regeneron Pharmaceuticals, Inc. (Tarrytown, USA). Medical writing assistance was provided by Richard J. Hogan, PhD, and Vojislav Pejović, PhD, of Eloquent Scientific Solutions, a division of Envision Pharma Group, and editorial and graphics assistance was provided by Eloquent Scientific Solutions. This support was funded by Sanofi.

