## Supplemental Appendix for "Sarilumab treatment of hospitalised patients with severe or critical COVID-19: a multinational, randomised, adaptive, phase 3, double-blind, placebo-controlled trial"

### Supplementary appendix

#### Figure legends

***Figure S1:*** **Kaplan-Meier curves of time to ≥2-point clinical improvement on the 7-point ordinal scale in patients with (A) severe and (B) critical disease, survival in patients with (C) severe and (D) critical disease, and time to discharge due to recovery in patients with (E) severe and (F) critical disease.**

***Figure S2:*** **Sarilumab concentration, pharmacodynamic markers, and laboratory findings potentially related to COVID-19 severity, over time.** (A) Mean (SD) serum sarilumab concentration by treatment and number of doses received. (B) CRP concentration. (C) Median IL-6 concentration. (D) sIL-6R concentration. (E) Neutrophil count. (F) Neutrophil to lymphocyte ratio. (G) D-dimer concentration. (H) ALT concentration. Data are mean (SE), except where otherwise indicated. ALT=alanine aminotransferase. COVID-19=coronavirus disease 19. CRP=C-reactive protein. IL=interleukin. sIL-6R=soluble IL-6 receptor.

***Figure S3:*** **Frequencies of initiations over time and by treatment group.** (A) Systemic corticosteroids. (B) Dexamethasone. (C) Antiviral agents. (D) Hydroxychloroquine/chloroquine. (E) Systemic antibacterial agents.

***Figure S4:*** **Proportion of patients with selected medication use over calendar time.** CQ=chloroquine; CS=corticosteroid; HCQ=hydroxychloroquine.

*Table S1:* **Summary of patients by country.**

| | **Placebo (n=84)** | **Sarilumab 200 mg (n=159)** | **Sarilumab 400 mg (n=173)** |
|---|---|---|---|
| Argentina | 3 (3·6) | 5 (3·1) | 6 (3·5) |
| Brazil | 13 (15·5) | 27 (17·0) | 37 (21·4) |
| Canada | 4 (4·8) | 8 (5·0) | 8 (4·6) |
| Chile | 16 (19·0) | 22 (13·8) | 21 (12·1) |
| France | 8 (9·5) | 20 (12·6) | 19 (11·0) |
| Germany | 1 (1·2) | 3 (1·9) | 2 (1·2) |
| Israel | 3 (3·6) | 2 (1·3) | 2 (1·2) |
| Italy | 6 (7·1) | 8 (5·0) | 11 (6·4) |
| Japan | 2 (2·4) | 1 (0·6) | 3 (1·7) |
| Russian Federation | 21 (25·0) | 47 (29·6) | 44 (25·4) |
| Spain | 7 (8·3) | 16 (10·1) | 20 (11·6) |

Data are n (%).

| *Table S2:* **Summary of selected medication use.** | | | | |
|---|---|---|---|---|
| | **Placebo** | **Sarilumab 200 mg (n=159)** | **Sarilumab 400 mg (n=173)** | **All (N=416)** |
| Antiviral agents | | | | |
| Prior use* | 15 (17·9) | 39 (24·5) | 40 (23·1) | 94 (22·6) |
| Prior and concomitant use† | 13 (15·5) | 39 (24·5) | 38 (22·0) | 90 (21·6) |
| Concomitant use‡ | 17 (20·2) | 49 (30·8) | 45 (26·0) | 111 (26·7) |
| Antibacterial agents | | | | |
| Prior use | 52 (61·9) | 101 (63·5) | 121 (69·9) | 274 (65·9) |
| Prior and concomitant use | 51 (60·7) | 97 (61·0) | 118 (68·2) | 266 (63·9) |
| Concomitant use | 70 (83·3) | 127 (79·9) | 147 (85·0) | 344 (82·7) |
| Azithromycin | | | | |
| Prior use | 29 (34·5) | 59 (37·1) | 78 (45·1) | 166 (39·9) |
| Prior and concomitant use | 26 (31·0) | 54 (34·0) | 72 (41·6) | 152 (36·5) |
| Concomitant use | 39 (46·4) | 73 (45·9) | 90 (52·0) | 202 (48·6) |
| Hydroxychloroquine/chloroquine | | | | |
| Prior use | 24 (28·6) | 55 (34·6) | 54 (31·2) | 133 (32·0) |
| Prior and concomitant use | 24 (28·6) | 54 (34·0) | 52 (30·1) | 130 (31·3) |
| Concomitant use | 29 (34·5) | 69 (43·4) | 65 (37·6) | 163 (39·2) |
| Systemic corticosteroids use | | | | |
| Prior use | 16 (19·0) | 25 (15·7) | 42 (24·3) | 83 (20·0) |
| Prior and concomitant use | 15 (17·9) | 24 (15·1) | 40 (23·1) | 79 (19·0) |
| Concomitant use | 39 (46·4) | 58 (36·5) | 78 (45·1) | 175 (42·1) |
| Dexamethasone | | | | |
| Prior use | 13 (15·5) | 11 (6·9) | 23 (13·3) | 47 (11·3) |
| Prior and concomitant use | 13 (15·5) | 10 (6·3) | 22 (12·7) | 45 (10·8) |
| Concomitant use | 23 (27·4) | 25 (15·7) | 42 (24·3) | 90 (21·6) |

Data are n (%).

*Before first infusion.

† Before and during/after first infusion.

‡During and/or after first infusion.

| *Table S3:* **Other secondary endpoints (mITT population) (day 60 analysis).** | | | |
|---|---|---|---|
| | **Placebo (n=84)** | **Sarilumab 200 mg (n=159)** | **Sarilumab 400 mg (n=173)** |
| **Analysis of proportion of patients alive at day 60** | | | |
| Patients alive at day 60, n (%) | 75 (89·3) | 142 (89·3) | 155 (89·6) |
| Difference *vs* placebo (95% CI)* | | 0·0 (−8·2 to 8·2) | 0·3 (−7·7 to 8·3) |
| p value *vs* placebo† | | 0·99 | 0·81 |
| **Analysis of time to resolution of fever‡** | | | |
| Kaplan-Meier estimates, days | | | |
| Median (95% CI)§ | 7·0 (6·0–12·0) | 8·0 (7·0–9·0) | 9·0 (7·0–10·0) |
| p value¶ | | 0·67 | 0·76 |
| Hazard ratio *vs* placebo (95% CI)# | | 0·91 (0·59–1·40) | 0·92 (0·60–1·40) |
| **Analysis of number of days with fever by day 29 for patients alive by day 29** | | | |
| LS mean (SE)** | 1·8 (0·3) | 1·2 (0·2) | 1·3 (0·2) |
| Difference *vs* placebo (95% CI) | | −0·6 (−1·4 to 0·2) | −0·4 (−1·2 to 0·3) |
| p value | | 0·08 | 0·21 |
| **Analysis of percent of days with fever by day 29** | | | |
| LS mean (SE)** | 12·7 (1·6) | 6·8 (1·2) | 8·0 (1·1) |
| Difference *vs* placebo (95% CI) | | −5·9 (−10·1 to −1·7) | −4·7 (−8·8 to −0·6) |
| p value | | 0·002 | 0·013 |
| **Analysis of time to NEWS2 of <2 and maintained for 24 hours** | | | |
| Kaplan-Meier estimates, days | | | |
| Median (95% CI)§ | 11·0 (8·0–14·0) | 9·0 (7·0–10·0) | 9·0 (8·0–11·0) |
| p value¶ | | 0·82 | 0·59 |

| | | | |
|---|---|---|---|
| Hazard ratio *vs* placebo (95% CI)# | | 1·05 (0·77–1·44) | 1·09 (0·80–1·48) |
| **Analysis of time to resolution of fever and improvement in oxygenation‡ ††** | | | |
| Kaplan-Meier estimates, days | | | |
| Median (95% CI)§ | 8·0 (7·0–12·0) | 9·0 (8·0–10·0) | 10·0 (9·0–13·0) |
| p value¶ | | 0·44 | 0·43 |
| Hazard ratio *vs* placebo (95% CI)# | | 0·82 (0·52–1·28) | 0·82 (0·53–1·26) |
| **Analysis of time to improvement in oxygenation††** | | | |
| Kaplan-Meier estimates, days | | | |
| Median (95% CI)§ | 7·0 (5·0–8·0) | 6·0 (5·0–7·0) | 6·0 (5·0–7·0) |
| p value¶ | | 0·29 | 0·57 |
| Hazard ratio *vs* placebo (95% CI)# | | 1·17 (0·86–1·58) | 1·11 (0·83–1·50) |
| **Analysis of proportion of patients alive off supplemental oxygen at day 29** | | | |
| Patients alive off supplemental oxygen at day 29, n (%) | 73 (86·9%) | 135 (84·9%) | 145 (83·8%) |
| Difference *vs* placebo (95% CI)* | | −2·0 (−11·1 to 7·1) | −3·1 (−12·2 to 6·0) |
| p value *vs* placebo† | | 0·61 | 0·63 |
| **Analysis of percent of days with events related to oxygen status by day 29‡‡** | | | |
| Percent of days with hypoxaemia | | | |
| LS mean (SE)** | 76·3 (2·7) | 73·0 (2·1) | 75·1 (1·9) |
| Difference *vs* placebo (95% CI) | | −3·3 (−10·4 to 3·8) | −1·2 (−8·2 to 5·8) |
| p value | | 0·31 | 0·70 |
| Percent of days with supplemental oxygen use | | | |
| LS mean (SE)** | 73·2 (2·7) | 70·6 (2·1) | 73·3 (2·0) |
| Difference *vs* placebo (95% CI) | | −2·7 (−9·8 to 4·5) | 0·1 (−7·0 to 7·1) |
| p value | | 0·42 | 0·98 |
| Percent of days with resting respiratory rate > 24 breaths/min | | | |

| | | | |
|---|---|---|---|
| LS mean (SE)** | 15·7 (2·1) | 14·7 (1·6) | 14·6 (1·5) |
| Difference *vs* placebo (95% CI) | | −1·01 (−6·5 to 4·5) | −1·2 (−6·6 to 4·3) |
| p value | | 0·69 | 0·64 |
| Percent of ventilator-free days in the first 28 days | | | |
| LS mean (SE)** | 77·3 (3·0) | 74·8 (2·2) | 75·7 (2·1) |
| Difference *vs* placebo (95% CI) | | −2·4 (−10·2 to 5·3) | −1·6 (−9·2 to 6·1) |
| p value | | 0·49 | 0·65 |
| **Number of days of events related to oxygen status for patients alive by day 29‡‡** | | | |
| Number of days with hypoxaemia, median (IQR) | 8·0 (5·0–14·0) | 7·0 (4·0–14·0) | 7·0 (5·0–14·0) |
| Number of days with supplemental oxygen use, median (IQR) | 8·0 (5·0–14·0) | 7·0 (4·0–13·0) | 7·0 (5·0–14·0) |
| Number of days with resting respiratory rate >24 breaths/min, median (IQR) | 0·0 (0·0–2·0) | 0·0 (0·0–2·0) | 0·0 (0·0–2·0) |
| Number of ventilator-free days in the first 28 days, median (IQR) | 29·0 (28·0–29·0) | 29·0 (28·0–29·0) | 29·0 (26·0–29·0) |
| **Analysis of time to saturation ≥94% on room air** | | | |
| Kaplan-Meier estimates, days | | | |
| Median (95% CI)§ | 8·0 (7·0–11·0) | 8·0 (6·0–10·0) | 8·0 (8·0–11·0) |
| p value¶ | | 0·70 | 0·80 |
| Hazard ratio *vs* placebo (95% CI)# | | 1·1 (0·8–1·4) | 1·1 (0·8–1·4) |
| **Initiation of mechanical ventilation, noninvasive ventilation, or use of high-flow nasal cannula** | | | |
| Patients with initiation of ventilation§§, n (%) | 13 (19·1) | 26 (20·5) | 33 (23·4) |
| Difference *vs* placebo (95% CI)* | | 1·4 (−10·3 to 13·0) | 4·3 (−7·4 to 16·0) |
| p value *vs* placebo† | | 0·81 | 0·61 |
| **Analysis of proportion of patients requiring rescue medication during the 28-day period¶¶** | | | |
| Patients requiring rescue medication, n (%) | 19 (22·6) | 22 (13·8) | 26 (15·0) |
| Difference *vs* placebo (95% CI)* | | −8·8 (−19·2 to 1·6) | −7·6 (−18·0 to 2·8) |

| | | | |
|---|---|---|---|
| p value *vs* placebo† | | 0·048 | 0·10 |
| **Analysis of proportion of patients with need for ICU care during study – Patients not in an ICU at baseline##** | | | |
| Patients with need for ICU care, n (%) | 7 (12·5) | 11 (11·2) | 17 (14·9) |
| Difference *vs* placebo (95% CI)* | | −1·3 (−12·0 to 9·4) | 2·4 (−8·4 to 13·3) |
| p value *vs* placebo† | | 0·98 | 0·54 |
| **Summary of number of days of hospitalisation among patients alive at day 60*** ** | | | |
| LS mean (SE)** | 15·9 (1·3) | 15·6 (1·0) | 16·1 (0·9) |
| Difference *vs* placebo (95% CI) | | −0·2 (−3·5 to 3·0) | 0·2 (−3·0 to 3·5) |
| p value | | 0·87 | 0·87 |
| **Summary of proportion of patients discharged due to recovery, n (%)** | | | |
| Day 4 | 3 (3·6) | 3 (1·9) | 6 (3·5) |
| Day 7 | 14 (16·7) | 33 (20·8) | 38 (22·0) |
| Day 15 | 43 (51·2) | 92 (57·9) | 102 (59·0) |
| Day 21 | 66 (78·6) | 113 (71·1) | 122 (70·5) |
| Day 29 | 70 (83·3) | 126 (79·2) | 137 (79·2) |
| Day 60 | 73 (86·9) | 135 (84·9) | 144 (83·2) |
| **Time from first dose to discharge due to recovery** | | | |
| **Kaplan-Meier estimates, days** | | | |
| Median | 14·0 | 11·0 | 13·0 |
| (95% CI)§ | (11·0–16·0) | (10·0–15·0) | (10·0–15·0) |
| p value *vs* placebo¶ | | 0·69 | 0·94 |
| Hazard ratio *vs* placebo | | 1·05 | 1·00 |
| (95% CI)# | | (0·79 to 1·40) | (0·76 to 1·33) |

*Based on asymptotic confidence limits.

†p value based on Cochran-Mantel-Haenszel test stratified by severity of illness (severe, critical) and use of systemic corticosteroids (Yes, No) as entered in the IRT.

‡Resolution of fever was defined as body temperature ≤36·6°C (axilla) or ≤37·2°C (oral) or ≤37·8°C (rectal or auricular) for ≥48 hours without antipyretics or until discharge. Patients who did have fever at baseline were censored at the first dose date.

§Two-sided 95% CI is computed by Brookmeyer and Crowley method (log-log transformation).

¶p value based on log-rank test stratified by severity of illness (severe, critical) and use of systemic corticosteroids (Yes, No) as entered in the IRT.

#Cox proportional hazard model stratified by severity of illness (severe, critical) and use of systemic corticosteroids (Yes, No) as entered in the IRT.

**The analysis of the endpoint was performed using the ANCOVA model with treatment group and randomisation strata as fixed effects.

††Improvement in oxygenation was defined as increase in $SpO_2/FiO_2$ of ≥50 compared with the nadir $SpO_2/FiO_2$ for ≥48 hours.

‡‡Resting respiratory rate maximum value was used if recorded at more than once a day. Hypoxaemia was defined as $SpO_2$ <93% on room air, or requiring supplemental oxygen or mechanical ventilatory support. Only the days meeting the criteria since the first dose were counted.

§§Mechanical ventilation, noninvasive ventilation, or use of high-flow nasal cannula.

¶¶Rescue medications were defined as the immunosuppressive therapies, as reported by investigator as “rescue therapy” on the Medications form.

##Even when ICU was not available.

***Days of hospitalisation since the first dose were counted.

ANCOVA=analysis of covariance. $FiO_2$=fraction of inspired oxygen. ICU=intensive care unit. IRT=interactive response technology. LS=least squares.

NEWS2=National Early Warning Score 2. $SpO_2$=oxygen saturation.

**Figure S1. Kaplan-Meier curves of time to ≥2-point clinical improvement on the 7-point ordinal scale in patients with severe (A) and critical disease (B), survival in patients with severe (C) and critical disease (D), and time to discharge due to recovery in patients with severe (E) and critical disease (F)**

### Figure S1A

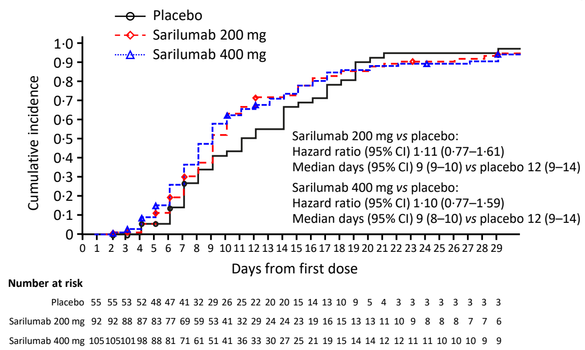


### Figure S1B

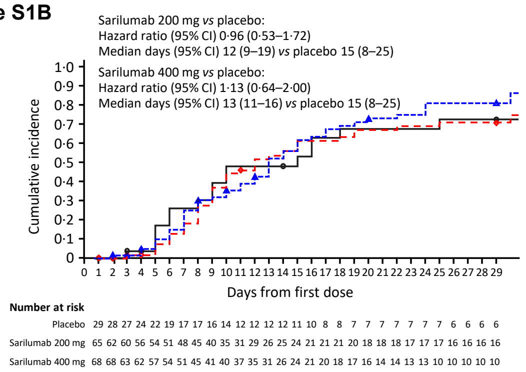


### Figure S1C

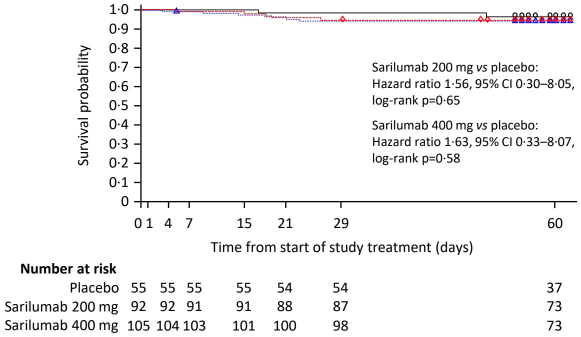


### Figure S1D

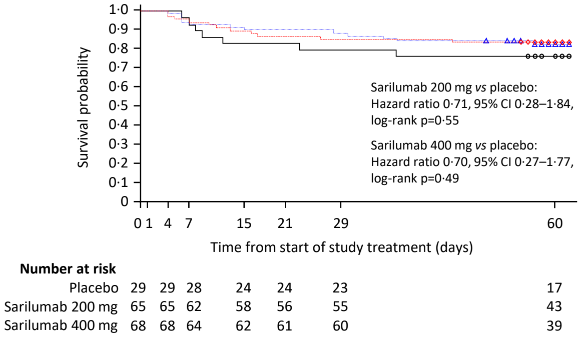


### Figure S1E

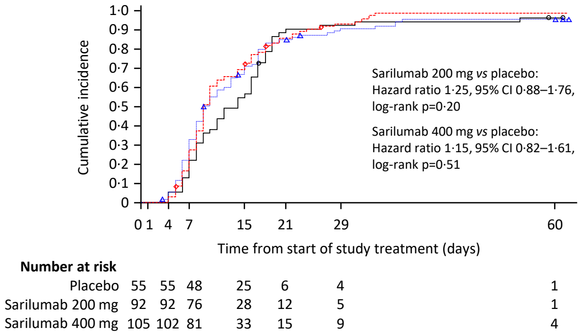


### Figure S1F

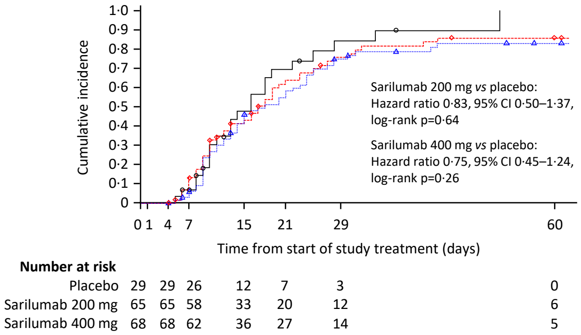

**Figure S2: Sarilumab concentration, pharmacodynamic markers, and laboratory findings potentially related to COVID-19 severity, over time**

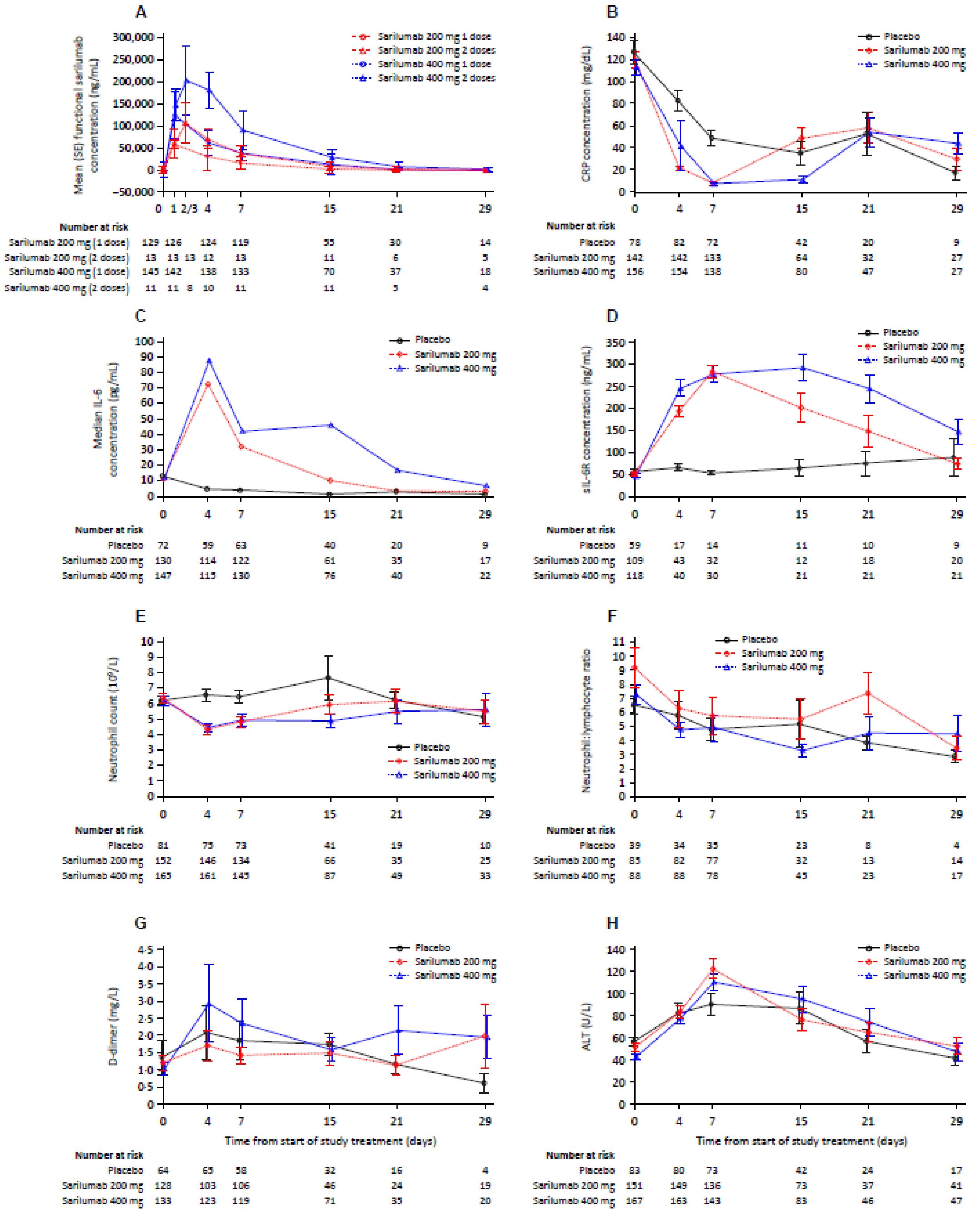

**Figure S3. Proportions of patients with initiation or continued use of systemic corticosteroids (A), dexamethasone (B), antiviral agents (C), hydroxychloroquine/chloroquine (D), and systemic antibacterial agents (E)**

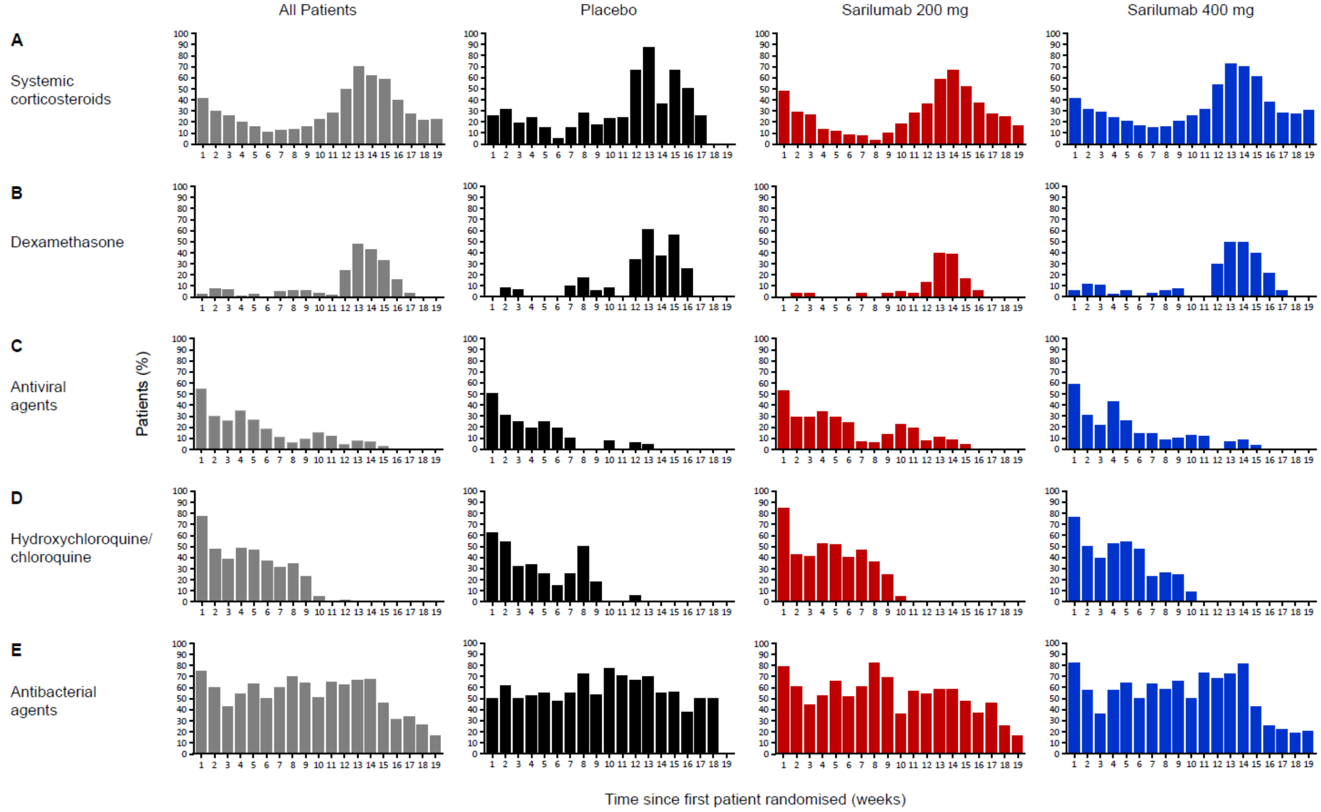

**Figure S4. Proportions of patients with selected medication use over time**

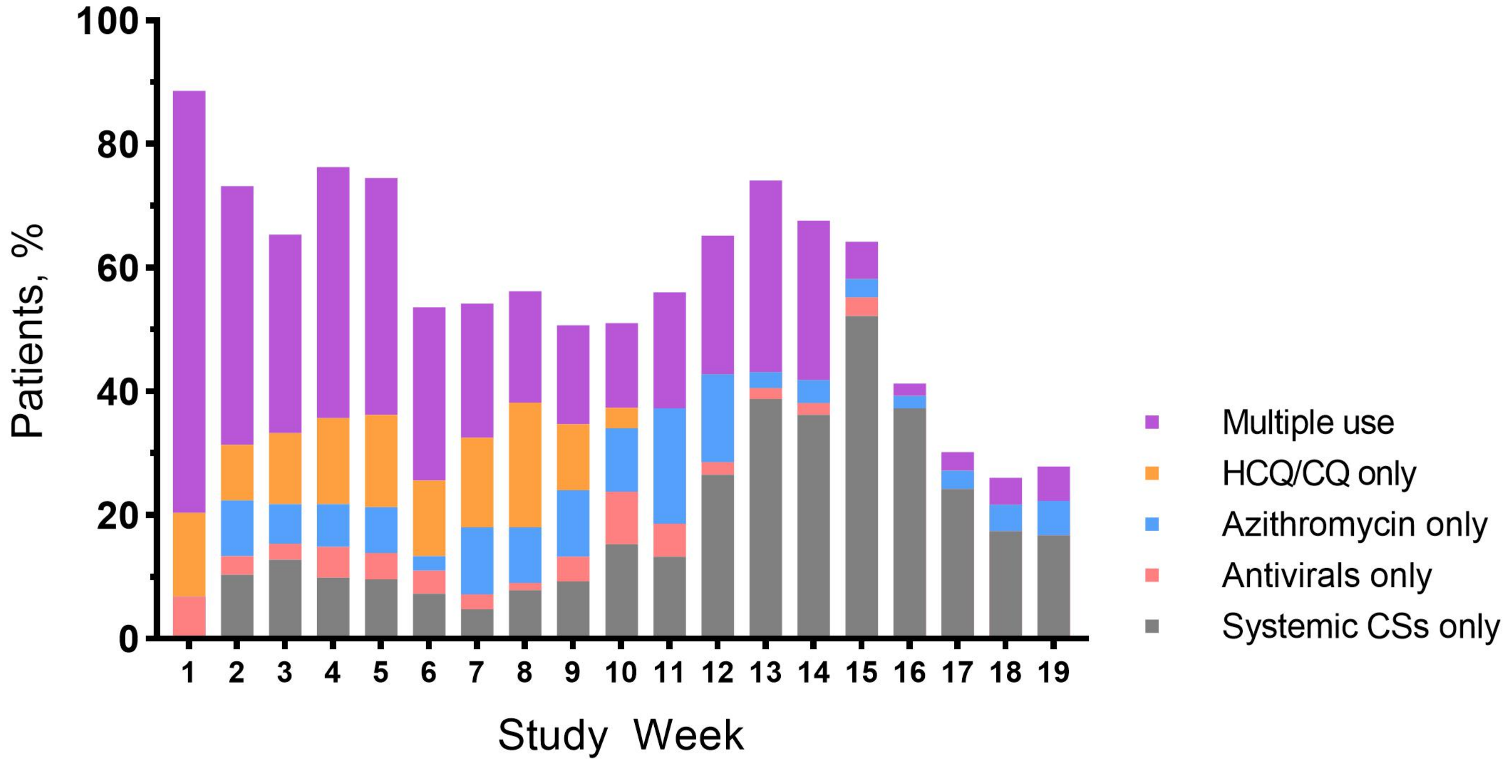
